# Quality improvement in emergency units in low resource settings: A qualitative assessment for understanding challenges and delivering solutions

**DOI:** 10.64898/2026.01.09.26343812

**Authors:** Indrakantha Welgama, Daniel Lange Da Silva, Gerard O’ Reilly, Aurore Nishimwe, Agnieszka Ignatowicz, Justine Davies

## Abstract

**Objective:** To meet the World Health Organization (WHO) requirement of strengthening integrated Emergency, Critical and Operative (ECO) care, it is necessary to develop ECO-related guidance and tools for Quality Improvement (QI) in Emergency Units (EUs). This study aimed to identify the barriers and facilitators for QI in EUs in Low- or Middle-Income- countries (LMICs) and explore potential measures for assessing quality of care, to inform guidance and tool development across all quality domains.

**Methods:** We conducted in-depth interviews (IDIs) with 11 specialist emergency physicians from LMICs and organized a workshop with global technical experts in emergency care. Transcribed interviews were thematically analysed, integrating WHO quality domains and the WHO Health Systems Framework. The workshop included presentations, discussions, and interactive ranking exercises to evaluate QI measurement methods and identify priority WHO quality domains. Related outcomes were synthesized to generate insights for the development of QI guidance and tools.

**Findings:** QI efforts in LMICs primarily focused on effectiveness, timeliness, safety, and occasionally patient centeredness. Key barriers to QI specific to LMIC EUs included high patient turnover, a lack of skilled staff, limited funds and resources, and prioritizing patient care quantity over quality. Facilitators included the availability of QI guidance, training, initiating small-scale projects, enhancing documentation and communication, and supportive hospital management. Safety and effectiveness were considered the most feasible QI domains for improvement, though all were recognized as important. Inconsistencies in how participants defined the domains highlighted the need to develop clear guidance on definitions, and importance of all domains and measures appropriate to each.

**Conclusions:** Emergency unit is a unique environment handling time-critical health conditions and therefore requires the development of specific tools and guidance for delivery of QI. Addressing barriers and leveraging facilitators identified in this study can inform the development of effective QI strategies for LMIC EUs.

## Introduction

Low- or middle-income countries (LMICs) encounter a large burden of emergency conditions (1–3), contributed to by a high incidence of injuries, infectious diseases, and complications of non-communicable diseases (2). Median Disability Affected Life Years (DALYs) attributed to emergency conditions are greatest in low-income countries followed by middle-income countries (4). Prior studies have identified that approximately 40% of deaths from injuries and other time-critical conditions are avoidable; with 60% of these avoidable deaths occurring due to issues in receiving quality care in health facilities (5, 6). In LMICs, similar to high income countries (HICs), patients with emergency conditions often present to Emergency Units (EUs) (7), where mortality is known to be greater than in HICs (1, 8–10). These findings for emergency conditions align with the findings of Lancet Global Health Commission on High Quality Health Systems, which suggests that poor quality of care in facilities is a substantial driver of adverse health outcomes (1).

Quality health care is experienced across multiple domains, as defined by the Institute of Medicine (IoM) (11) and updated by the World Health Organization (WHO) (12). These quality domains include clinical effectiveness, safety, people-centeredness, equity, efficiency, timeliness, and integrated care, and the WHO has described them as follows (12).

- Effective: “*providing evidence-based healthcare services to those who need them*”.
- Safe: “*avoiding harm to people for whom the care is intended*”.
- People centred: “*providing care that responds to individual preferences, needs and values*”.
- Equitable: “*providing care that does not vary in quality on account of gender, ethnicity, geographic location, and socio-economic status*”.
- Efficient: “*maximizing the benefit of available resources and avoiding waste*”.
- Timely: “*reducing waiting times and sometimes harmful delays*”
- Integrated care: “*providing care that makes available the full range of health services throughout the life course*”.

Quality health care is essential for successful universal health coverage (13), and guidance to assist providers with improving quality of care is developed by WHO (14). However, despite efforts to promote quality improvement (QI) initiatives, their adoption and implementation remains challenging in LMICs (15, 16). Deficiencies in multiple WHO Health Systems Framework building blocks (17), currently experienced by most LMICs, including the lack of skilled healthcare providers, under-developed information technology and data collection, poor access to essential medicines and equipment, conspire with financial constraints and governance issues to impede QI efforts, lead to adverse outcomes (18–21). As per the Donabedian framework (22), these deficiencies relate to either the “structure” and/or the “process”, which ultimately affects the “outcome” as lowered quality of care to the patients. These issues are magnified in EUs, which are unique environments with specific QI needs related to a large number of patients with a heterogenous mix of conditions of varying degrees of severity, and requiring time-critical care, being attended to in relatively brief encounters (23).

World Health Assembly resolution 76.2 of 2023 on “*Integrated emergency, critical and operative (ECO) care for universal health coverage and protection from health emergencies*” was ratified in response to challenges affecting patients in need of emergency care (24). Subsequently, the WHO has been requested to develop a global strategy and action plan to assist in the implementation of this resolution (25, 26). QI initiatives within the EUs will be central in achieving resolution 76.2, since EUs receive most Emergency, Critical and Operative (ECO) conditions presenting to health facilities. Furthermore, implementing QI initiatives in EUs is essential, as these settings should prioritize timeliness, safety, and clinical effectiveness through standardized protocols and efforts to enhance system-wide efficiency (18, 27). However, there is limited understanding of the specific challenges that impede QI delivery in EUs or how best to address these, particularly in LMICs.

To inform the development of Emergency, Critical and Operative care (ECO care) related guidance and tools for QI delivery in EUs in LMICs, this study aimed to identify, from a WHO quality domains perspective, the barriers and facilitators healthcare workers in EUs encounter when implementing and delivering programmes. We further aimed to identify measures for assessing quality of care within each WHO quality domain and determine which domains should be prioritized for tool and guidance development.

## Methods

This study was conducted in two phases. First, we conducted a qualitative study consisting of in-depth interviews (IDIs) to explore barriers and facilitators affecting QI delivery in EUs in LMICs. Second, we hosted an expert workshop to examine and reflect on these barriers and facilitators, assess methods for measuring the WHO QI domains, and identify which domains should be prioritised for development of guidance for QI in EUs.

### Phase 1: Qualitative interviews

#### Study setting

EUs in LMICs that had attempted to implement or maintain QI initiatives. LMICs were defined as countries whose Gross National Income (GNI) per capita was less than US $14,005, as per the World Bank country classification for 2024 (28).

#### Participants

Emergency care physicians with current or recent experience of QI initiatives in LMICs were selected as study participants. Potential participants were purposively selected by the WHO’s technical team on ECO care. The selection process leveraged WHO’s existing knowledge of these physicians’ experience to ensure the inclusion of individuals with relevant insights into QI implementation in EUs.

#### Data collection

IDIs were conducted by IW and DL between October 2023 and February 2024. A Topic Guide (TG) was developed based on findings from previous studies (29, 30) and authors’ knowledge of delivery of QI and providing emergency care (Supplementary Material 01). Questions in the TG enquired about current QI practices in EUs and the barriers and facilitators in implementing such activities. The TG was piloted prior to use. All IDIs were conducted virtually and recorded for post interview transcription. Interviews were conducted in English, given participants were proficient in the language. Data collection and analysis were undertaken concurrently such that the IDIs continued until thematic saturation was reached (31).

#### Data analysis

Audio recordings were transcribed by IW and DL and the transcripts were uploaded to NVivo software (32) for coding and analysis. Interviews were analysed thematically by IW, DL, AI and JD, following the approach described by Braun and Clarke (33) and incorporating elements of the constant comparative method (34). Initially, a sample of interview transcripts was reviewed to generate inductive codes, focusing on barriers and facilitators to QI initiatives. Deductive coding was also applied based on the WHO quality domains (12) and the WHO Health Systems Framework (17). Thereafter, all transcripts were coded against this framework, with further refinements made through discussion among authors to ensure consistency and accuracy. Findings were summarized quantitatively, reporting the number of participants per code, and qualitatively, using illustrative quotes to provide deeper insights.

#### Ethics

Ethics clearance was obtained from the Ethics Review Board of the University of Birmingham, United Kingdom (ERN_1742-Oct2023). Participation in the study was voluntary, and informed consent was obtained from all participants.

### Phase 2: Workshop with technical experts

#### Workshop setting

In Geneva, Switzerland.

#### Participants

Clinical and academic emergency physicians, along with other healthcare professionals involved in EU care, were invited to participate in the workshop. Participants, representing different countries (with different income levels and geographical regions), were purposefully selected by the WHO technical team on ECO care, based on the WHO’s prior experience, and ensuring participants’ knowledge and experience of delivering QI in LMIC settings.

#### Data collection and analysis

The meeting was structured around plenary presentations and discussions. Findings from the phase-1 qualitative study were presented, and participants were requested to reflect upon and articulate their experiences of barriers and facilitators for QI in EUs. This was followed by a presentation and discussion on the WHO quality domains, with questions posed on how the domains could be measured in the EUs, and responses captured as unlimited inputs using eMenti® (eMenti.com). Finally, after a presentation on QI processes in the EU and the current availability of WHO tools and guidance for QI (Supplementary Material 02), participants were requested to rank the WHO quality domains based on their perceived feasibility for change through QI interventions, using Slido® (Slido.com).

Outputs were captured through meeting notes, transcripts of recorded meeting proceedings, and responses from eMenti® and Slido®. Given that QI requires providers to identify, measure, and change exiting practices, and such change requires capability, opportunity, and motivation for change, the study findings were synthesised and summarized by the research team focusing on: (1) challenges of practicing QI in EU settings in LMICs; (2) potential facilitators to overcome these challenges; (3) methods for measuring WHO quality domains in the EUs; and (4) domains that are most feasible to change.

#### Ethics

Participation at the workshop was voluntary upon acceptance of the invitation, and verbal consent was obtained from all meeting attendees for using the meeting content and discussion points, while anonymity of the information was maintained. Furthermore, the final report of the workshop was shared with all workshop participants for their comments and confirmation. No patient or public involvement was required for this study.

## Results

Eleven IDIs were undertaken, and 26 participants attended the workshop (Table 1).

**Table 1.**
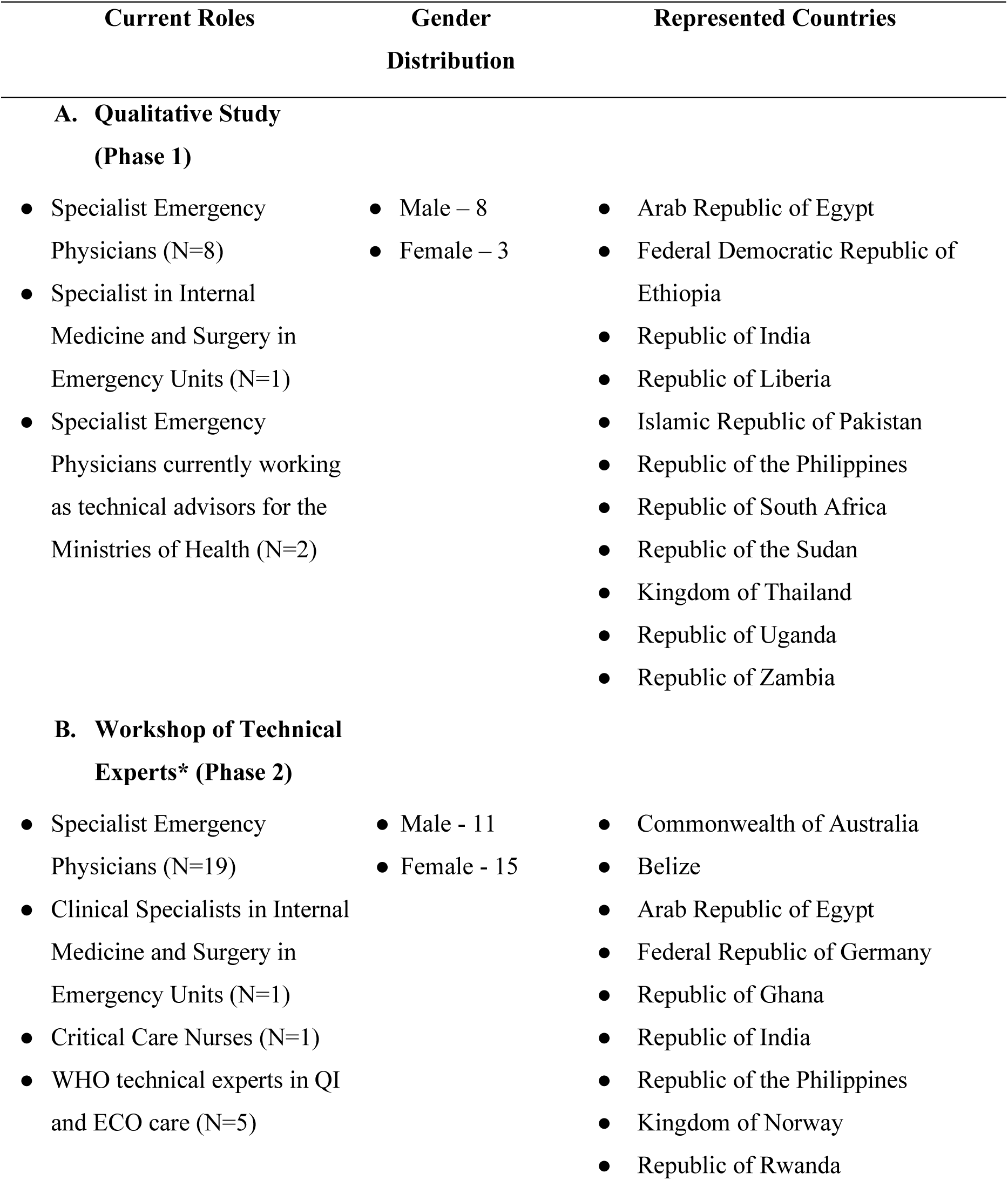
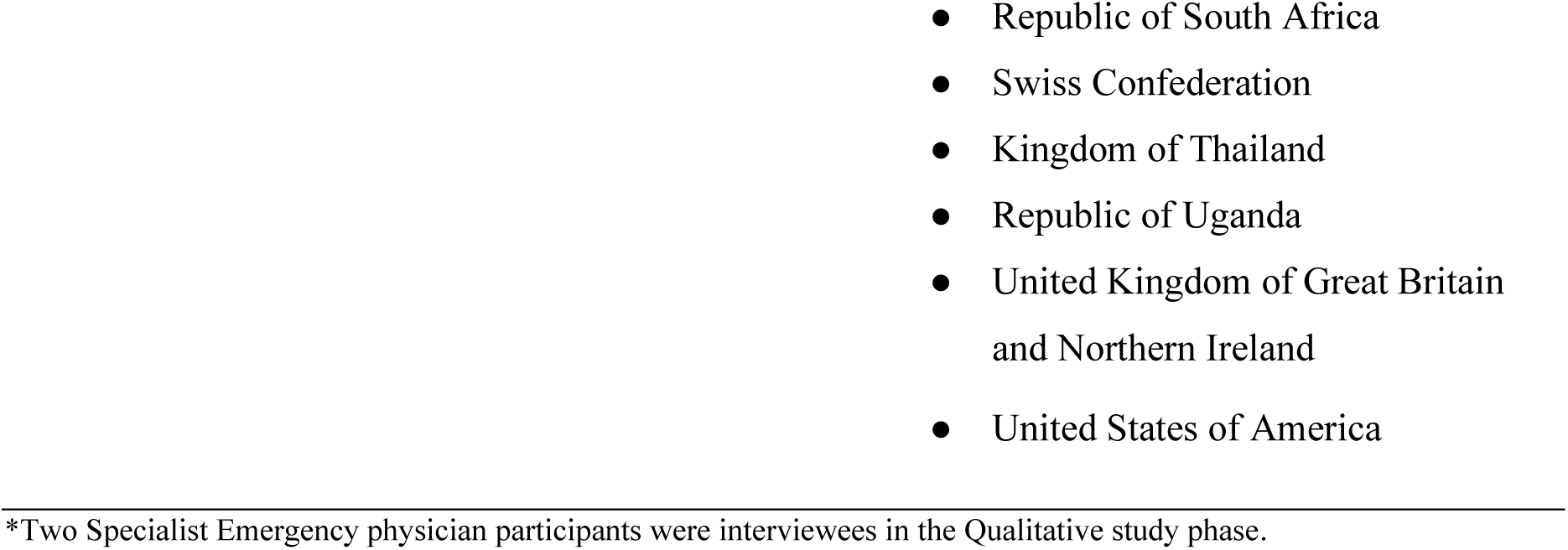
Participant characteristics in the qualitative interviews study and the workshop.

### Results of phase 1: Qualitative interviews study

All 11 IDI participants confirmed that QI activities were being conducted within their EUs. Amongst those activities, 25 specific longstanding initiatives were identified, primarily focused on improving clinical effectiveness, timeliness, safety, and person-centeredness. Five participants reported newly introduced QI initiatives, initiated by them. All 11 EUs had QI initiatives related to clinical effectiveness; none had initiatives focused on equity, or efficiency (Figure 1). Examples of these initiatives included reducing patient queues, initiating triage, staff training, obtaining patient feedback, conducting data-informed quality of care reviews, and routine monitoring using Key Performance Indicators (KPIs). Further details on QI initiatives are provided in Supplementary Material 03.

**Figure 1.**
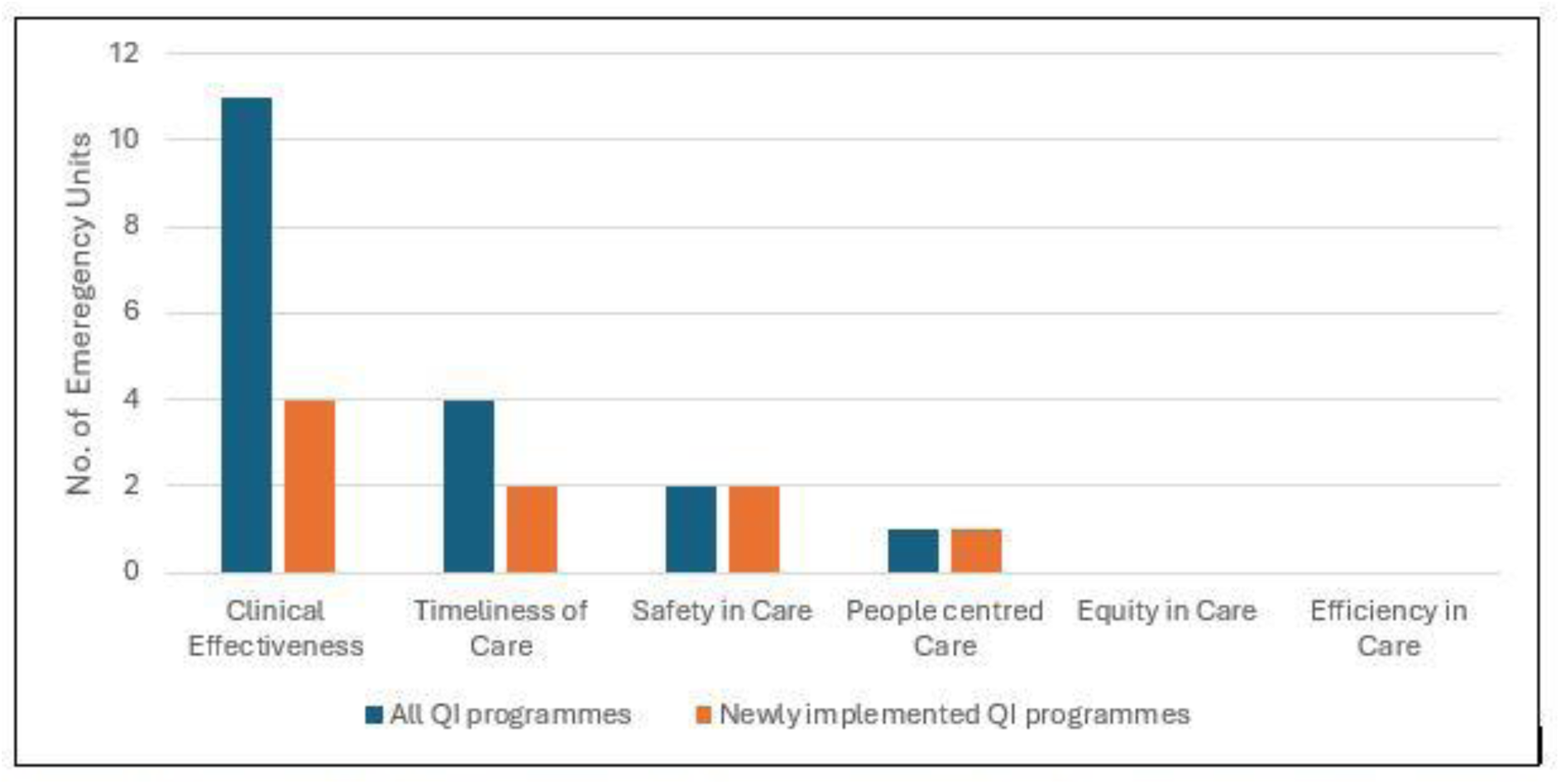
Number of EUs implementing QI Initiatives as per WHO Quality domains, and the number of EUs where new QI programmes were implemented by the participants (N=11).

#### Barriers to QI initiatives

Numerous barriers related to developing and implementing QI initiatives, categorised as per WHO Health Systems Framework (17) are presented in Table 2. The detailed findings, including issues specific to WHO QI domains are presented in Supplementary Material 04 - Section A.

**Table 2.**
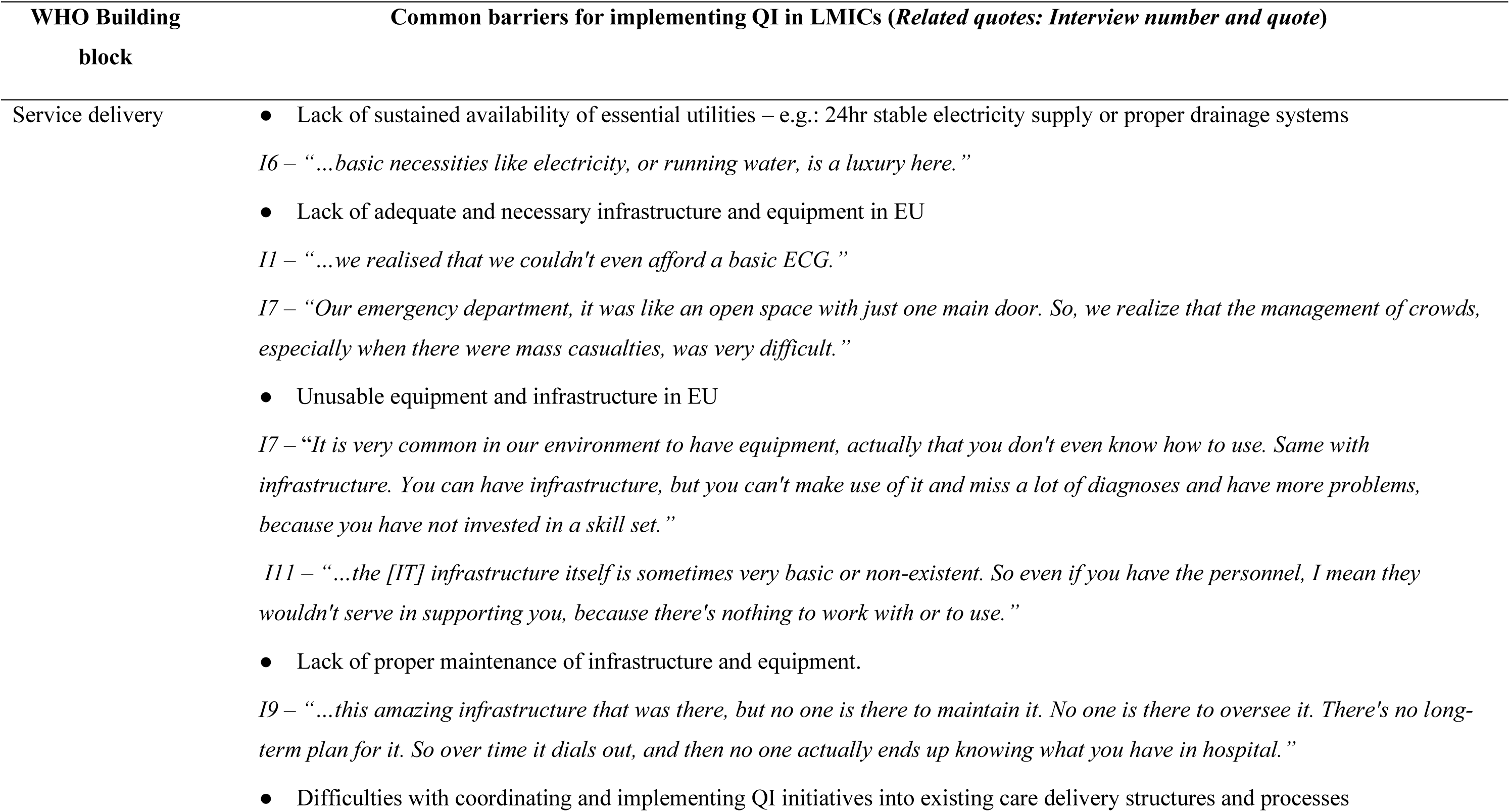
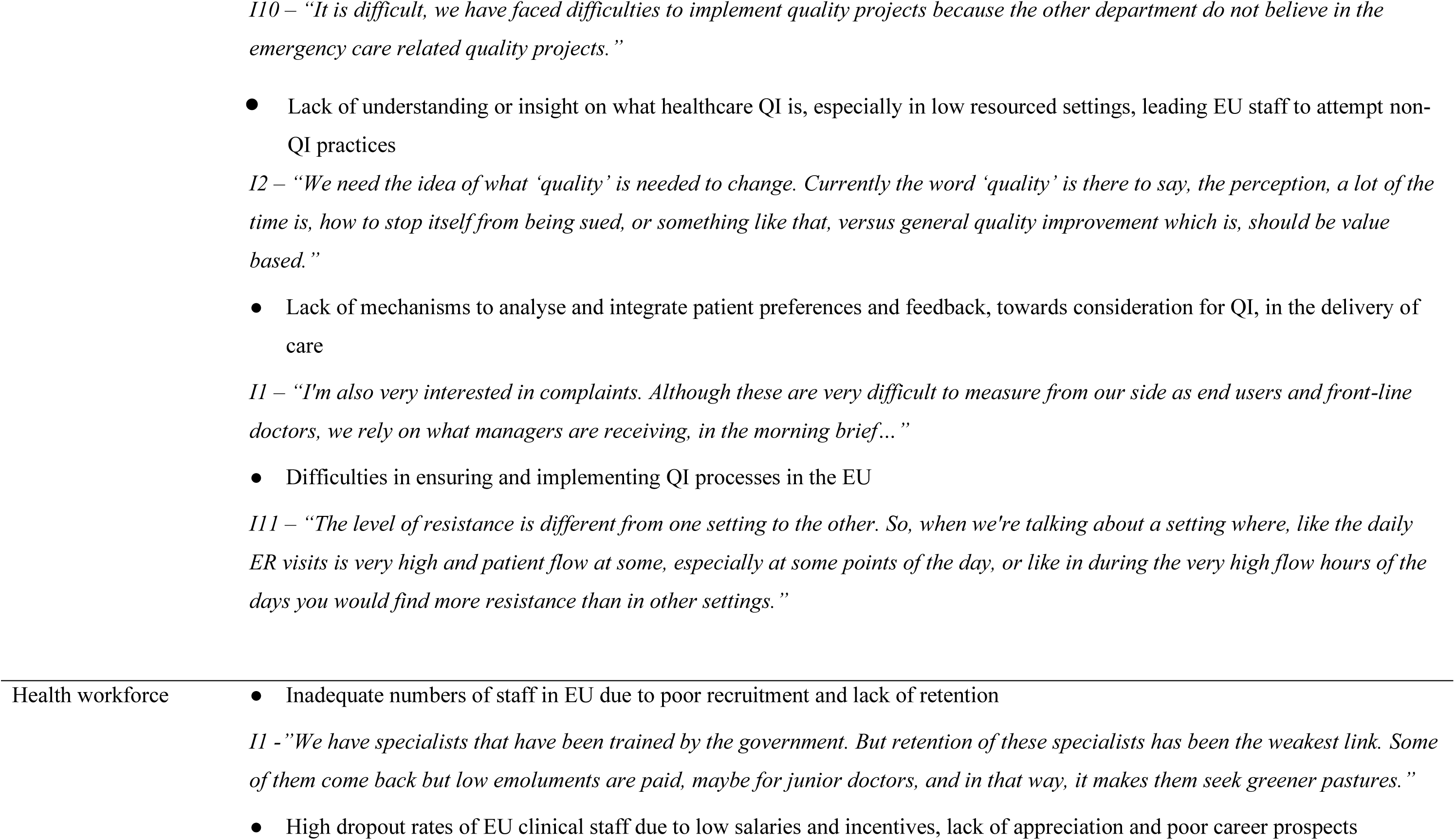
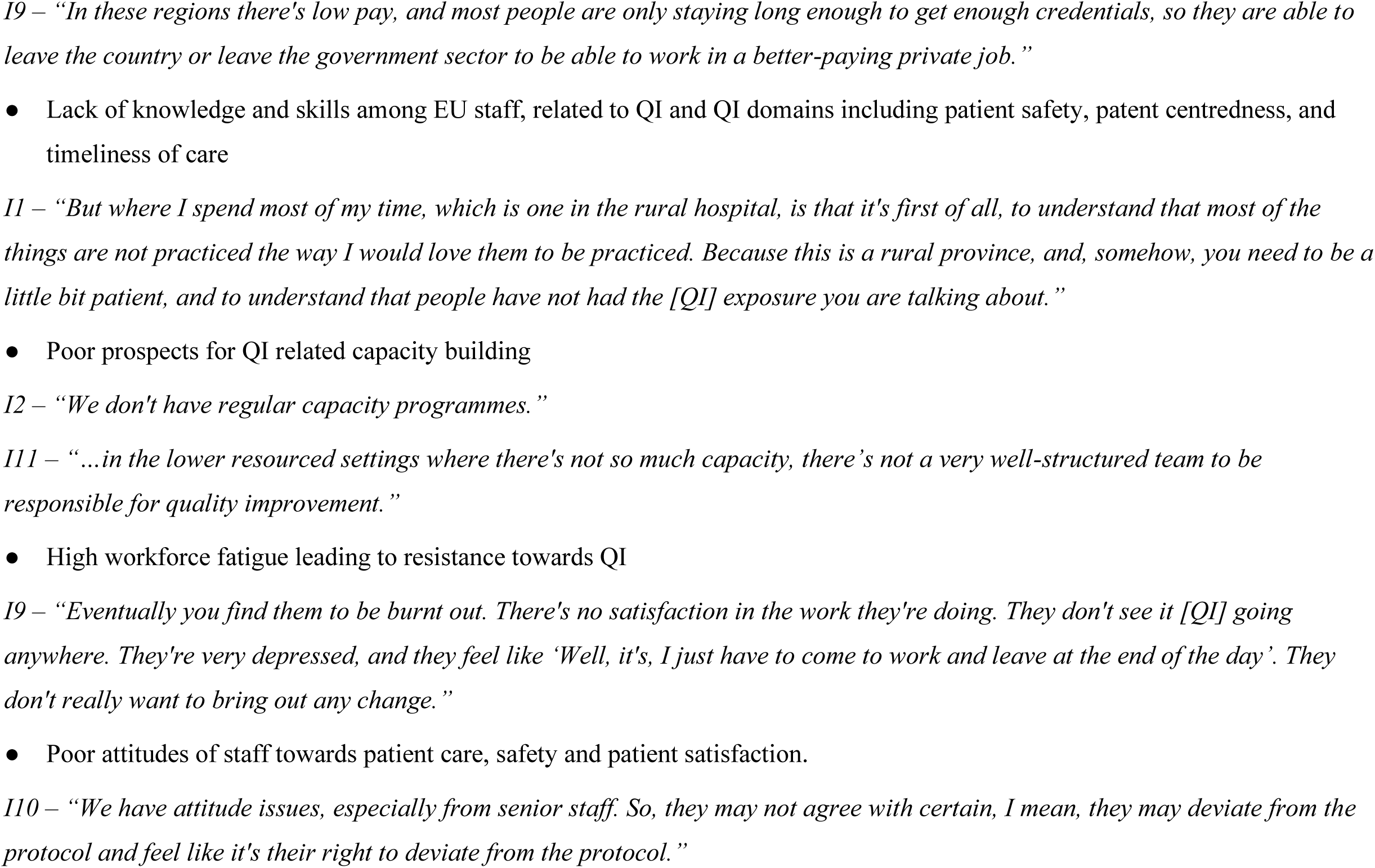
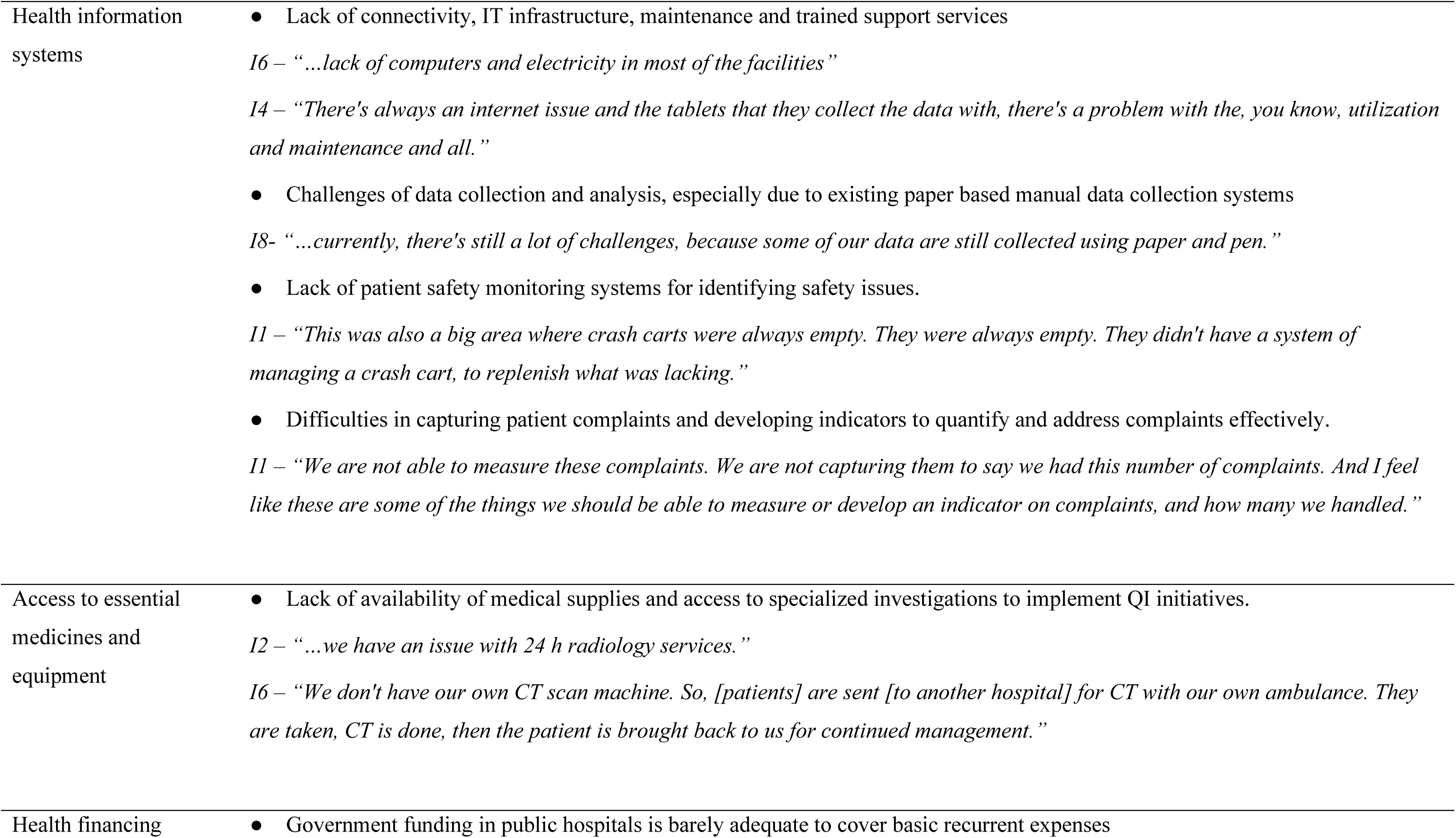
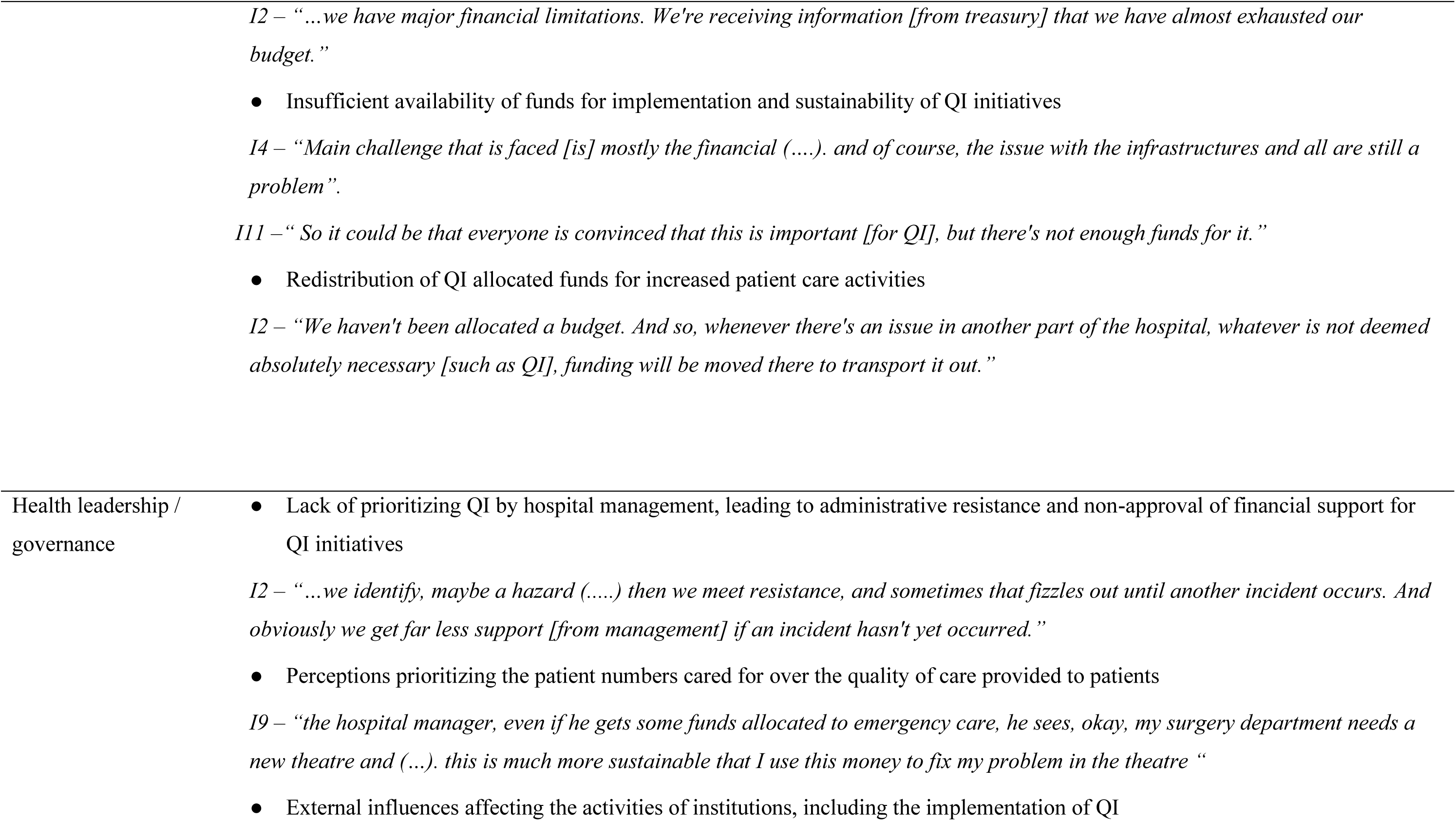
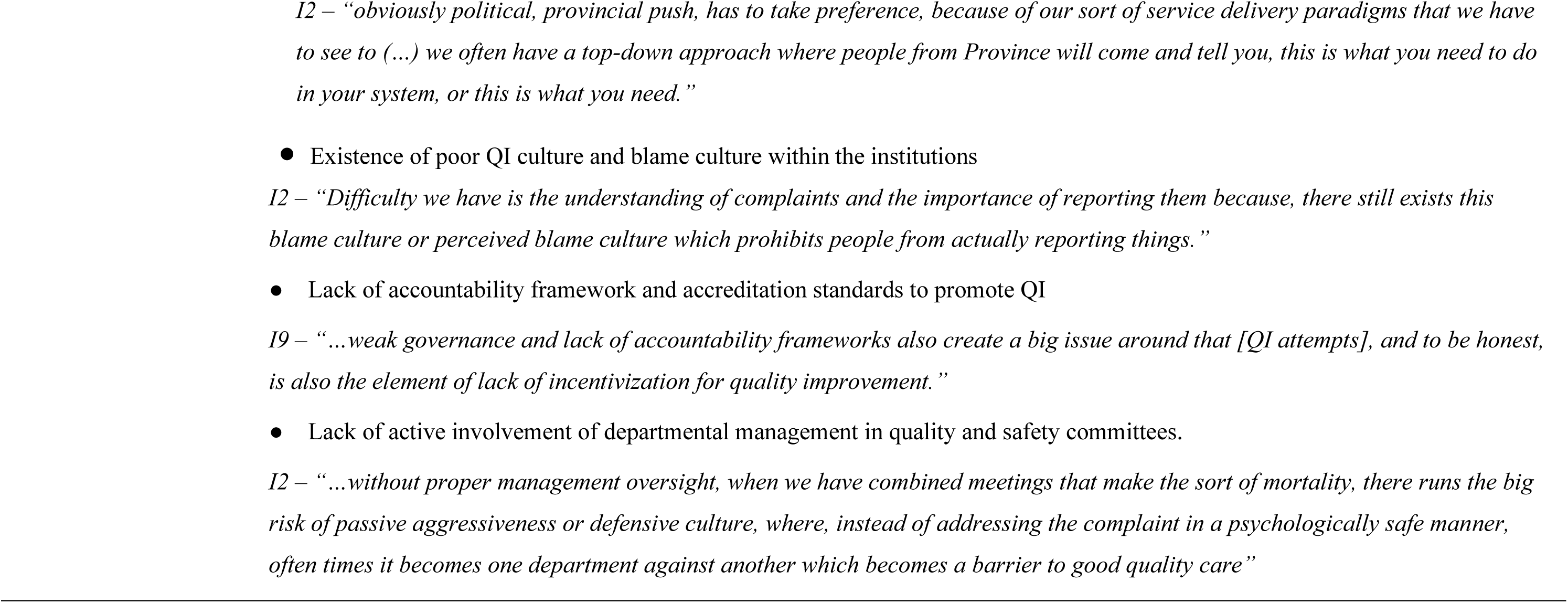
Barriers for Quality Improvement in EUs of LMICs.

Of note, when discussing barriers participants described challenges at different levels. Some explicitly referred to barriers that directly hindered QI delivery, while others highlighted broader challenges to delivering quality care which indirectly impeded ability to deliver QI. This highlights the complexity of separating challenges in delivering quality care from those specifically hindering QI implementation.

#### Facilitators for QI

Participants highlighted several factors which facilitated their attempts at implementing QI. These are summarised in Table 3, with details presented in Supplementary Material 04 - Section B.

**Table 3.**
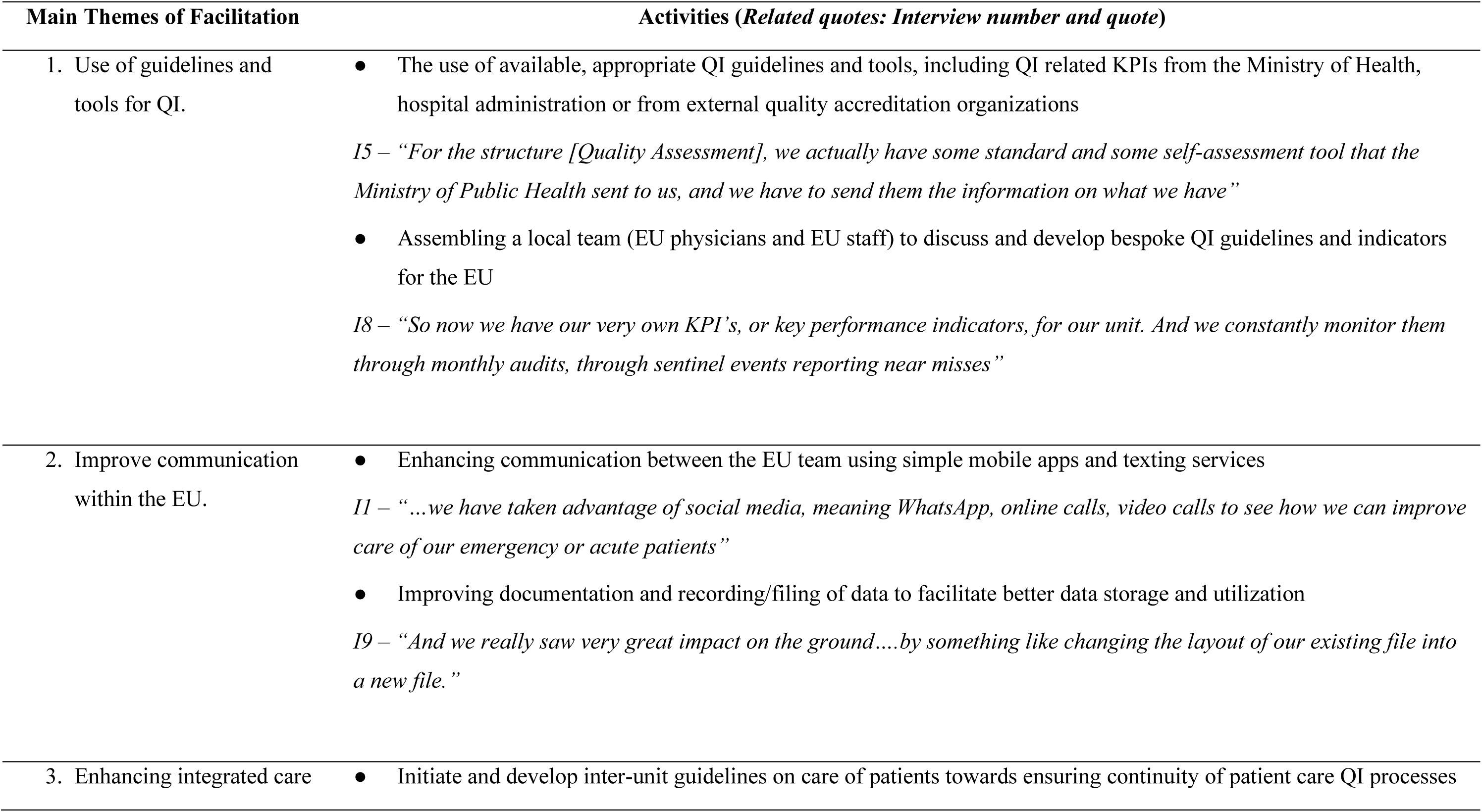
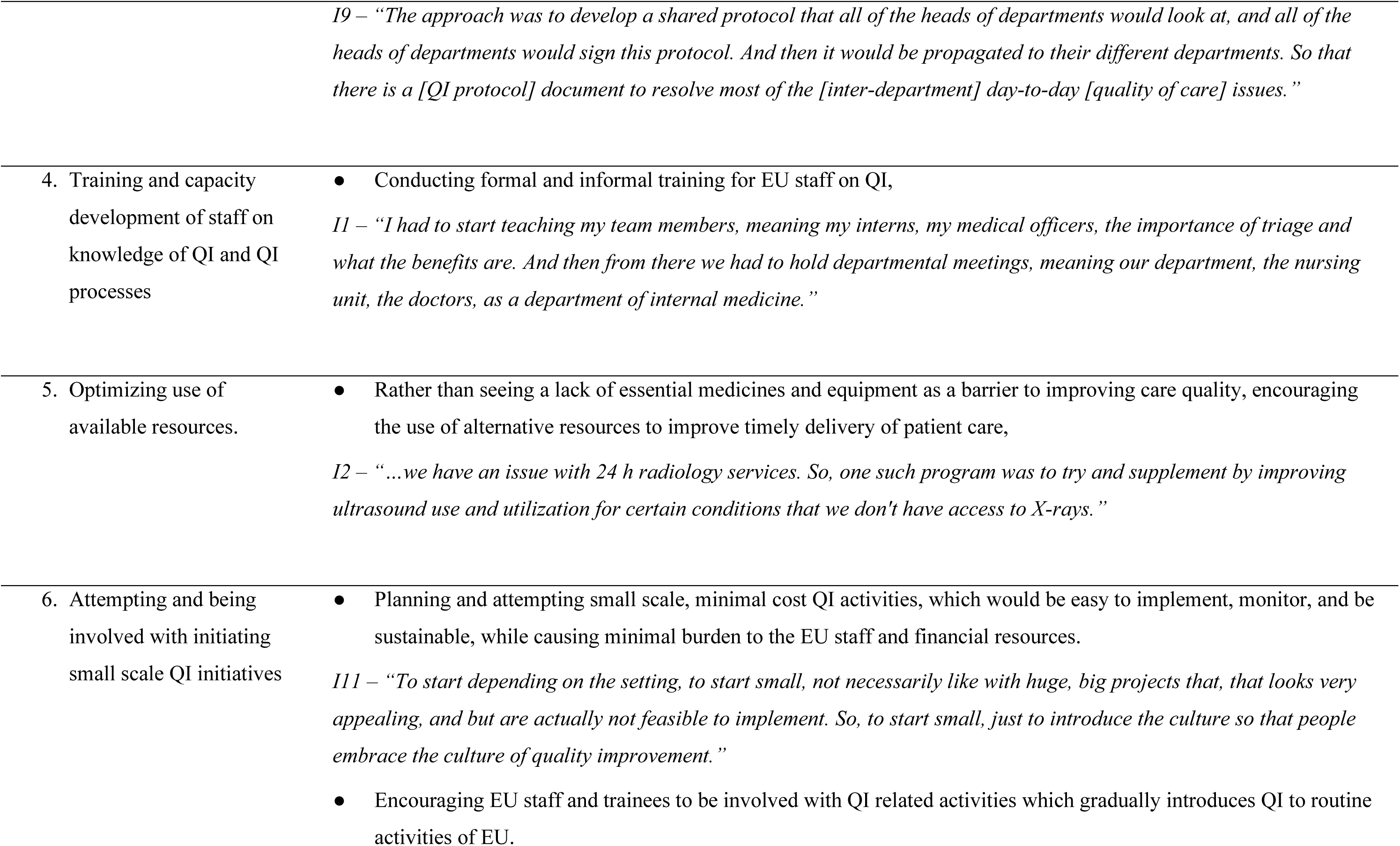
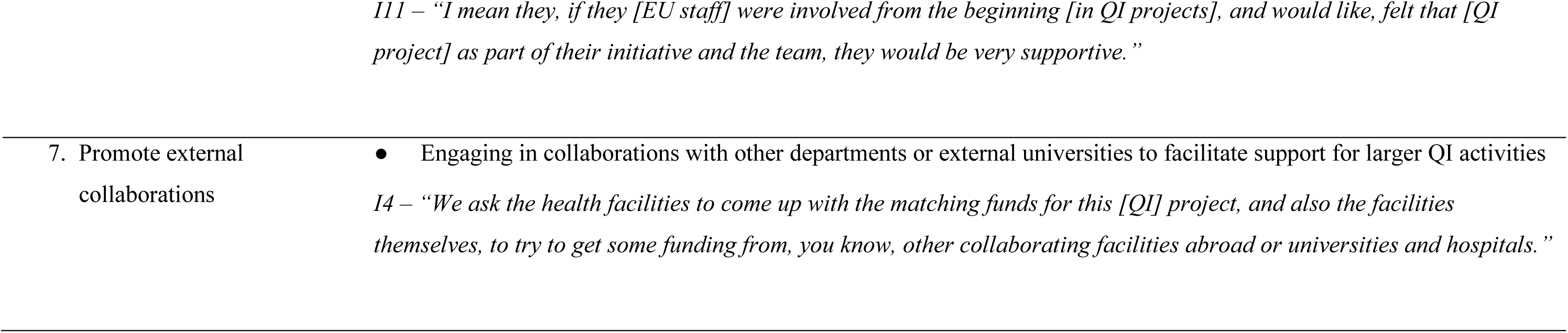
Suggested facilitators for QI in EU.

### Results of phase 2: Workshop with Technical experts

#### Suggestions for measuring quality domains in the EU

Summarised responses for how to measure WHO QI domains are presented in Table 4, with the word clouds and analysis of responses presented in the Supplementary Material 05. Results show that whilst some domains were clearly demarcated, the categorisations of others were blurred. For instance, timeliness of care was included as a measure to assess effectiveness of care, and measures for efficiency were offered under measures of timeliness. Some participants presented domain definitions rather than methods for measurement.

**Table 4.**
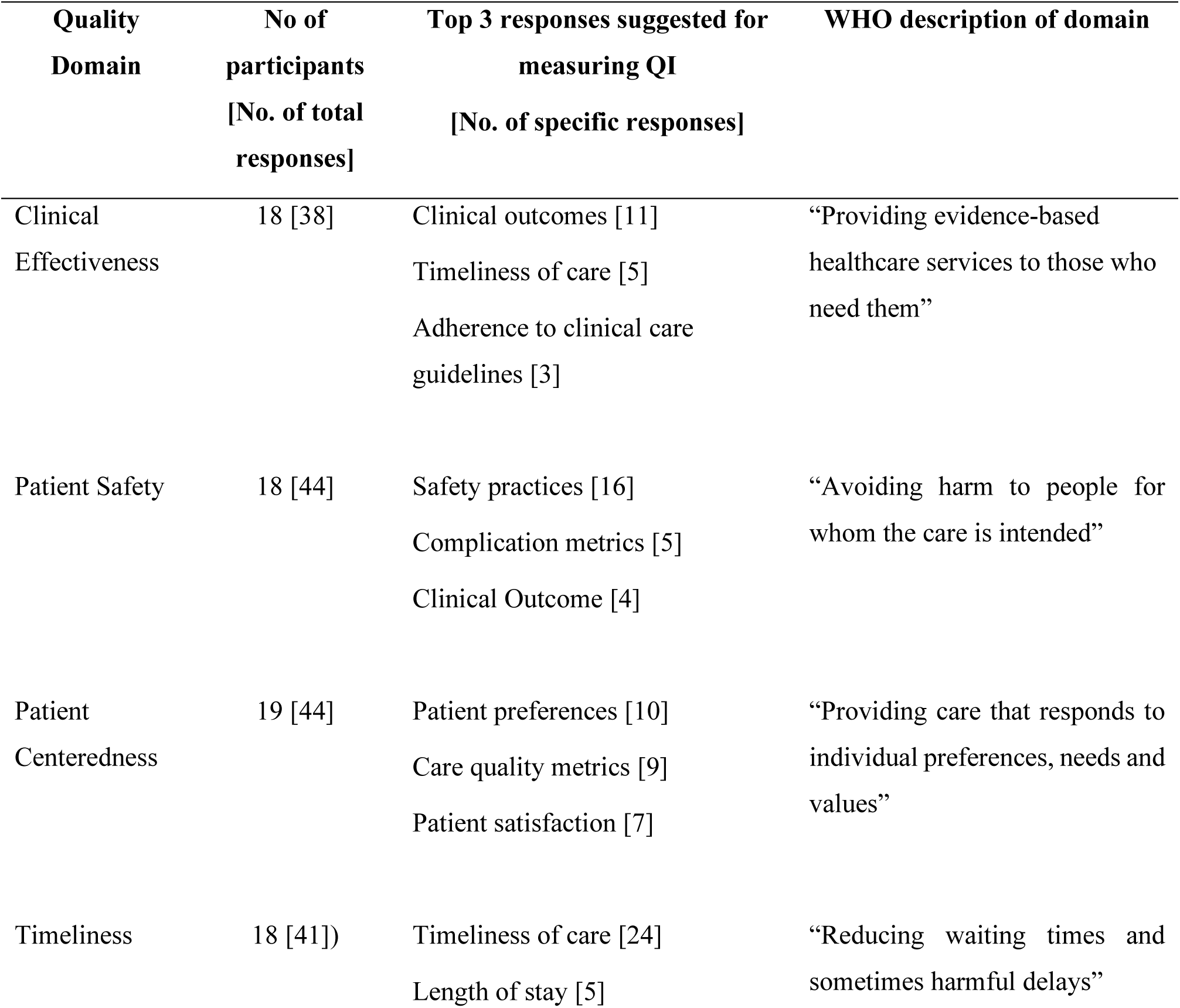
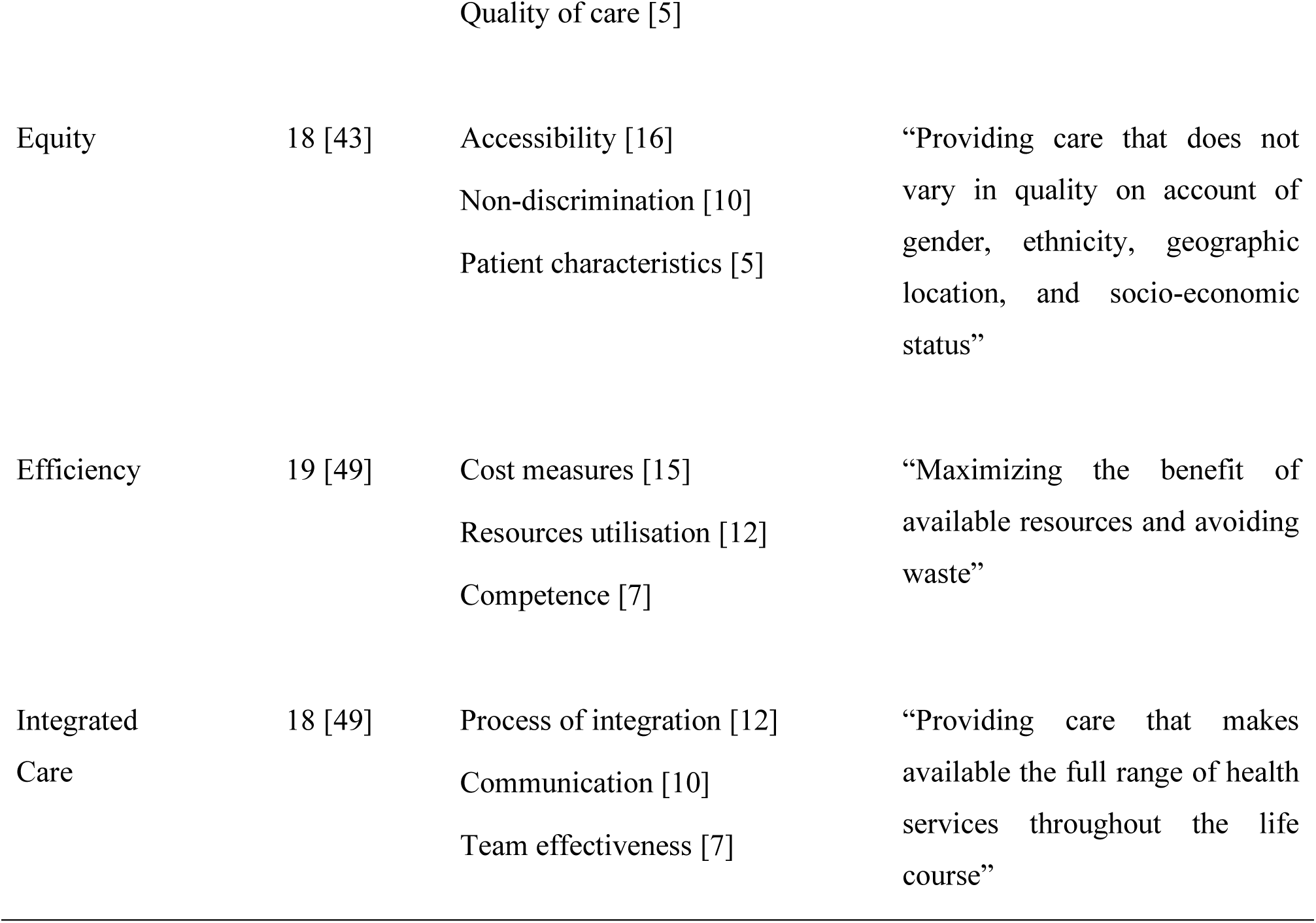
Key responses suggested by technical experts for measuring the concepts of WHO Quality domains.

#### Perceptions on feasibility of WHO Quality domains to change

Safety (76.5%) was ranked by the workshop participants, as the most feasible domain to change, followed by clinical effectiveness 52.9%. Patient centredness and timeliness of care were equally ranked (41.2%) as the third most feasible domain for change. Efficiency (29.4%), Integrated care (17.6%) and Equitable care (11.8%) were ranked lower in feasibility for change. None agreed that it would be feasible to change all quality domains simultaneously.

#### Barriers and facilitators to implementing QI in the EU settings

Reflecting on findings from the qualitative interviews, workshop participants posed that barriers were primarily related to three key factors. First, EUs face a heavy patient load, making it difficult to implement QI initiatives effectively. Second, patient diagnoses are often unknown during their stay in the EU, adding complexity to clinical decision-making. Third, the EU is not the final point of care for patients but is instead deeply interconnected with other hospital services and external healthcare interactions. Participants also emphasized the importance of frontline healthcare staff safety in EUs. Given the high-pressure environment, ensuring the personal safety of healthcare providers was highlighted as a critical concern.

Suggested facilitators were generic to QI delivered in other units, but felt to be of particular importance in EU, given the aforementioned challenges. For example, ensuring QI programmes are developed with inputs from staff and patients was recognized as a way to enhance local ownership and relevance of the QI programmes. Participants highlighted the need for broad, adaptable improvement strategies rather than interventions reliant on specific clinical care pathways, allowing for quality improvements regardless of case mix and diagnoses. Working towards a minimum standard of care was suggested as an EU-specific facilitator. However, other participants preferred to work towards improvements from baseline. Incorporating assessments of quality of care delivered prior to patients arrive in the EU was considered important, but difficult to capture. Finally, participants agreed that ensuring QI initiatives are feasible in resource constrained EUs was essential for the sustainability of QI. Detailed findings are presented in Supplementary Material 06.

## Discussion

This study explored the delivery and implementation of QI initiatives in EUs in LMICs, identifying barriers, facilitators, and feasibility considerations for QI activities. Our findings provide insights into existing QI efforts and implementation challenges, emphasizing the need for structured, adaptable tools and guidance to support and sustain QI efforts in EUs.

The interview data revealed that existing QI programmes primarily focused on clinical effectiveness, safety, and timeliness of care, aligning with findings of prior studies (35–38) . However, patient-centeredness was less frequently targeted, while equity and efficiency were absent, highlighting gaps in QI efforts. Clinical effectiveness was the most frequently implemented QI initiative, yet barriers such as limited guidelines, lack of timely and accurate information, and resource constraints significantly hindered the impact. Workshop discussions emphasized the need for substitute resources in LMIC settings to mitigate these challenges, particularly in the context of staffing shortages and the absence of structured QI capacity-building programmes (39).

Workshop participants identified safety as the most feasible domain for improvement, reinforcing the global emphasis on patient safety in hospitals (14, 37, 40). They cited administrative, financial, and reputational concerns as key drivers for prioritizing patient safety, emphasizing the need for targeted guidance to strengthen QI implementation in EUs. While safety remained a top priority, participants acknowledged that all QI domains are interdependent and essential for delivering high-quality care (13). Discussions highlighted the challenge of balancing these priorities, particularly in resource-constrained settings, and underscored the need for a comprehensive approach to QI.

Patient-centeredness was ranked third in feasibility for change, with several measurement approaches suggested. Patient-reported measures are considered reliable indicators (41, 42), but workshop participants noted challenges in collecting and analysing patient complaints and service reviews, indicating a need for EU-specific measures of patient-centeredness. Timeliness of care was ranked equally feasible to change; it was also the third most common focus of QI initiatives among interview participants’ hospitals. Numerous timeliness-based Key Performance Indicators (KPI) were being used by interview participants as standardized measurement methods. The integrated nature of care in the EU, where services rely on other departments such as diagnostics and referral systems, emphasized the need for specific timeliness metrics tailored to emergency settings. Equity and efficiency were largely absent from QI efforts, in interviews with hospital providers despite equity’s recognized importance in universal health coverage (43). Whereas amongst workshop participants, efficiency was often misinterpreted with timeliness. Workshop participants faced challenges towards defining measurable indicators for integrated care, reinforcing the need for practical tools and measures to improve coordination within EUs.

Whilst none of the workshop participants thought that all quality domains were equally important, the overlapping definitions of WHO quality domains, particularly timeliness, effectiveness, and efficiency, suggests that they are difficult to disentangle. These findings mirror those from other healthcare settings where these concepts were often thought to be interconnected and difficult to distinguish clearly (36). The interdependency of the domains means that improvement in quality in any domain may have beneficial effects on others and knowledge of all domains is important to ensure that none are neglected (44). Lack of clarity around the quality domains suggests that future guidance should emphasize understanding all domains, ensuring that QI efforts do not neglect any. There should also be suggestions for measurements appropriate for each and all domains, and guidance should illustrate completing the QI loop, for multiple domains, broader than the usual ones of effectiveness and safety (45).

Our findings indicate that barriers to QI in EUs align with WHO’s Health Systems Framework (17), spanning infrastructure deficits, workforce limitations, governance weaknesses, and system-wide inefficiencies. Context-specific challenges in LMICs, such as unreliable electricity, inadequate infrastructure, and equipment shortages, underscore the need for QI tools that accommodate resource constraints while maintaining measurable improvements in care quality. Additionally, workforce shortages and a lack of QI training highlight the importance of developing accessible training materials, toolkits, and implementation frameworks to equip frontline healthcare workers with the skills needed to integrate QI into routine practice. While some of our findings align with prior studies (4, 15, 46), there is limited research exploring all aspects of QI implementation across all WHO quality domains.

Despite these barriers, several facilitators were identified that could enhance QI programme development and implementation. The use of QI guidelines and locally developed protocols, tailored to EU-specific workflows, was considered critical. This suggests that new guidance should build on existing frameworks while allowing for local customization, ensuring that QI tools remain practical, adaptable, and relevant to EU settings. Strengthening internal communication within EUs and across hospital departments was also highlighted by participants as an important strategy for integrating QI into broader healthcare systems. In resource-constrained settings, small-scale, resource-efficient QI initiatives were seen as effective starting points, allowing gradual implementation without overwhelming existing systems. Additionally, training and capacity building were emphasized as essential for embedding QI knowledge among EU staff, while external collaborations with universities, NGOs, and international organizations were suggested as ways to secure funding and technical support for sustainable QI implementation.

This study highlights the need for LMIC-specific guidance and tools to support QI implementation in EUs. The QI resources must address existing gaps, particularly in equity, efficiency, patient-centeredness and integrated care, while providing suggestions to overcome infrastructure, workforce, and governance-related challenges. The development of adaptable QI tools - including training programs and standardized implementation guides - will be essential in ensuring sustained improvements in emergency care quality across diverse LMIC settings.

Our study has several limitations, primarily related to the participant profile and geographic representation. Most participants were senior emergency physicians, which may have introduced bias in emphasizing certain QI domains over others, potentially shaping the prioritization of clinical effectiveness while underrepresenting perspectives on other aspects of QI. The IDIs primarily included participants from Africa and Asia, with no representation from the Pacific and America regions, potentially limiting the generalizability of findings to other LMIC settings. However, this limitation was partially mitigated by the broader geographic representation of participants in the expert workshop. While efforts were made to capture a range of perspectives, differences in health system structures, policy environments, and available resources may have influenced participants’ experiences with QI. Finally, because data were collected through semi-structured interviews and a workshop, findings are based on self-reported experiences and perceptions. Our participants were QI experts, and our data may not have captured the concerns of those less engaged in QI efforts in LMICs.

## Conclusion

EUs are a unique environment within health facilities, which encounter time-critical patients who require urgent, but effective and safe interventions for survival, often linked with other specialized units, while ensuring patient centred, efficient and equitable care. However, a clear restriction is observed within EUs in LMICs, where QI appears to focus mainly towards enhancing the effectiveness and timeliness of care. Despite the numerous barriers encountered by EUs, linked to internal factors within the EU, as well as external factors such as governance and finance, opportunities exist, supported by internal and external facilitators, to enhance care quality across all seven quality domains. Process enhancing activities requiring minimal resources and implementation time can have a meaningful impact as their cumulative effects lead to major improvements in care quality. However, practical measures and indicators are essential to monitor progress effectively and ensure that all QI domains are addressed. Such tools will enable a more systematic and balanced approach to QI, fostering sustainable improvements in emergency care delivery in low resourced settings.

## Funding

This work was supported by the Quality Related (QR) research funding 23/24, issued by Research England through the University of Birmingham [QR Funding for Policy Support Fund: PSF-10]. The funders were not involved in the study design, implementation, data collection and analysis, nor the interpretations and submission of the paper for publication.

### Patient Consent for Publication

Not applicable.

## Acknowledgement

Authors wish to acknowledge Lee Wallis, Vanessa Naidoo and Rose Mabelle Buhain of the Clinical Services and Systems Unit, World Health Organization, for their technical guidance.

## Data availability statement

Analysed data are available upon reasonable request.

