## Supplementary material for "Quality improvement in emergency units in low resource settings: A qualitative assessment for understanding challenges and delivering solutions"

Findings on the challenges and facilitators for quality improvement in emergency units from the workshop of technical experts – Phase 2 of the study

### Supplementary Material 01

#### Topic Guide for the Qualitative In-Depth Interview Study - Phase 1 of the study.

- *Good morning / afternoon.*
- *How are you today?*
- *Thank you very much for joining us today for this interview.*
- *I am …* (introduction of self by the interviewer) *….and I will be the person interviewing you today.*
- *I hope you were able to read the information sheet shared earlier with you, about this study and participating in the interview.*
- *At the onset, I would like to reaffirm your consent for participating in his study. Your consent will be recorded prior to the interview recording, but it will not be included into the data section or data analysis. For the consent section only, I will be stating your name and institution and inquiring if you give consent to participate in this study. The consent section will be over with your response.*
- *Thereafter it will be the interview proper where I will ask you to state your work role relevant to the potential or actual quality improvement (QI) initiatives. I would like to remind you to please **not** state your name or the name of the country or facility in which you work in the interview proper. Is that fine with you?*
- *Right, I will start the recording now.*

**Interviewer, please start the recording now.**

**Consent section:**

- *As per the information sheet shared with you, I will be asking you about your plans, needs, and readiness for QI in the emergency department.*
- *The discussion should last less than one hour.*
- *At whatever time, if you feel uncomfortable with the discussion or the questions, you are free to let me know about that, and you may not answer such questions.*
- *Can you confirm that you, (Interviewee name) from (Name of country) give your consent to take part in this interview?*

**Interviewer, please make sure that the participant has clearly stated that they consent/said 'yes' to the above question.**

- *Thank you and that concludes the consent section of the interview.*
- *Now, we shall start with the interview proper.*

***A. Basic Details of Respondent***

1. *Can you please describe your work-role, relevant to your involvement with actual or potential quality improvement programmes?*

***B. Quality Improvement programmes***

2. *Considering Quality Improvement programmes in general, do you currently have an active quality improvement programme in the emergency department in your hospital?*

**If NO, please skip to Q.12 in page 3.**

If **YES**, please continue:

3. *Can you tell me more about that programme and how it functions?*

   *Prompts:*

   *What are the aims of this programme?*

   *Prompts*

   *What aspects of health care is it intending to improve?*

   *For Patients – If so, for which types of patients*

   *For Infrastructure*

   *For processes*

   *Is there a champion or a motivated leader of your QI programme?*

   *Is there a QI team?*

   *If yes, who are its members (i.e. categories of staff)*

   *How well is it organised?*

4. *How is quality of care assessed?*

   *Prompts:*

   *What observations or data do you use to assess quality?*

   *Who performs the assessments for care-quality?*

5. *What happens when quality of care is found to be lower than desired?*

   *Prompts:*

   *Does anything change?*

   *How are these changes made?*

   *Who is in-charge of making changes?*

   *Is there any reassessment of whether the changes have worked?*

6. *Are there any quality improvement meetings?*

| If YES: | If NO: |
|---|---|
| • *How often are they?* | • *Do you think that QI meetings would be useful?* |
| • *Who attends these meetings?* | • *Who do you think should attend?* |
| • *Are they well attended?* | • *How often do you think they should be?* |
| • *Do you think that all people who need to attend them are there? (if not, why?)* | |

7. *Do you think that your quality improvement programme is well supported?*

   *Prompts:*

   *By colleagues in the ED and senior level?*

   *By Hospital administration?*

   *With funding?*

   *With training?*

8. *How do you think the quality improvement programme is perceived by the staff in the ED?*
9. *How do you think the quality improvement programme is perceived by hospital management?*
10. *What are the main challenges that you or your colleagues face in maintaining the QI programme?*
11. *Based on your experience, if you were going to set up a new quality improvement programme, how do you think you would do this?*

    *Prompts:*

    - *Who would need to be involved?*
    - *Would any resources be needed?*
    - *What steps would you need to go through to make this successful?*

**Please skip to Q.23 in Section C in page 5.**

The Following questions up to section C, need to be answered only if the respondent has <u>stated as "**NO**" to having an existing QI programme.</u>

12. *Do you think a QI programme would be useful for your ED and why (if yes or no)?*
13. *Do you think that your ED is ready to start a QI programme? (if yes or no, why?)*

14. *What do you think is needed for your ED to be ready to start a quality improvement programme?*

    *Prompts:*

    - *Who would need to be involved?*
    - *Would any resources be needed? (If so, what would they be?)*
    - *What steps would you need to go through to make this QI programme successful?*

15. *What do you think a quality improvement programme would be most useful for?*

    *Prompts:*

    - *For what aspects of care*
    - *For what type of patients*

16. *How do you think quality of care would best be assessed?*

    *Prompts:*

    - *What observations or data do you think would be useful to assess quality?*
    - *Who would do the assessments of quality?*

17. *What do you think would happen if the quality of care is found to be lower than desired?*

    *Prompts:*

    - *Would anything change?*
    - *Who do you think would be in charge of making changes? (QI team or few individuals?)*
    - *Do you think there would be any assessment of whether such changes have worked?*

18. *Do you think there would be a need for any "Quality improvement meetings"?*

| **If YES:** | **If NO:** |
|---|---|
| • *How often?* | • *Why not?* |
| • *Who would attend?* | |
| • *Do you think they would be well attended?* | |
| • *Do you think that all people who need to attend them would be there?* | |

19. *Do you think that the quality improvement programme would be well supported?*

    *Prompts:*

    - *By colleagues in the ED and senior level*
    - *With funding*
    - *With training*

20. *How do you think that a QI programme would be perceived by staff in the ED?*

21. *How do you think that a QI programme would be perceived by hospital management?*

22. *What are the main challenges that you or your colleagues would possibly face in maintaining the QI programme?*

**C. All participants – Use of registries for quality improvement**

23. *Do you currently use data from a registry to help with quality improvement?*

If **NO**, please **skip to Q.30** in the next page.

If **YES**, please continue.

24. *What sort of registry is used for this purpose?*
    *Prompt*
    - *Is it a manual register or an electronic registry?*
    - *Is it a bespoke registry or developed locally?*

25. *Did you encounter any challenges in the set up or maintenance of that registry?*

    If YES, what were the challenges you or your colleagues encountered?

    *Prompts*

    - *Any hesitancy from ED colleagues or hospital management?*
    - *How about resources, were these available as needed?*

      *Prompts*

      - *funding and financial resources*
      - *human resources*
      - *IT, technological and infrastructure requirements*

26. *How do you think that registry can be further improved?*

27. *Do you monitor the quality of data that your registry collected?*
    *If so, how did you do this?*

28. *If you encountered issues of quality, what did you do to improve this?*

29. *Do you think that the quality of the data collected was good enough for it to be used for decision making?*

**Please skip to section D: "Conclusion" section in page 6.**

The Following questions up to section D, need to be answered only if the respondent has stated as “NO” to Q.23 above, on the use of data from a registry.

30. *Do you think having a registry would be useful for data collection towards quality improvement?*

31. *Do you think there would be challenges in the set up or maintenance of a registry?*

    If YES, what challenges would you anticipate?

    *Prompts*

    - *Any hesitancy from ED colleagues or hospital management?*
    - *how about resource availability?*

      *Prompts*

      - *funding and financial resources*
      - *human resources*
      - *IT, technological and infrastructure requirements*

32. *Do you think data quality would be an issue in maintaining such a registry?*

    *If YES, how do you think this could be improved?*

**D. Conclusion**

*Thank you very much for joining this interview today.*

*It was very informative, and we were able to gather a lot of information on the actual or potential Quality Improvement initiatives in an emergency department. Your experiences and opinions on the readiness for Quality Improvement in emergency care, is highly valuable for this study.*

*This will be very useful when we compile the information and disseminate it, for other countries and hospitals who are intending to implement and maintain a quality improvement programme in their emergency care setting.*

*Thank you very much and wish you a good day.*

### Supplementary Material 02

#### Findings from search of currently available WHO resources for QI in the EU highlighting gaps and areas for improvement

We developed a linearized conceptualisation of the QI process, which consisted of 10 stages (Figure 2.1).

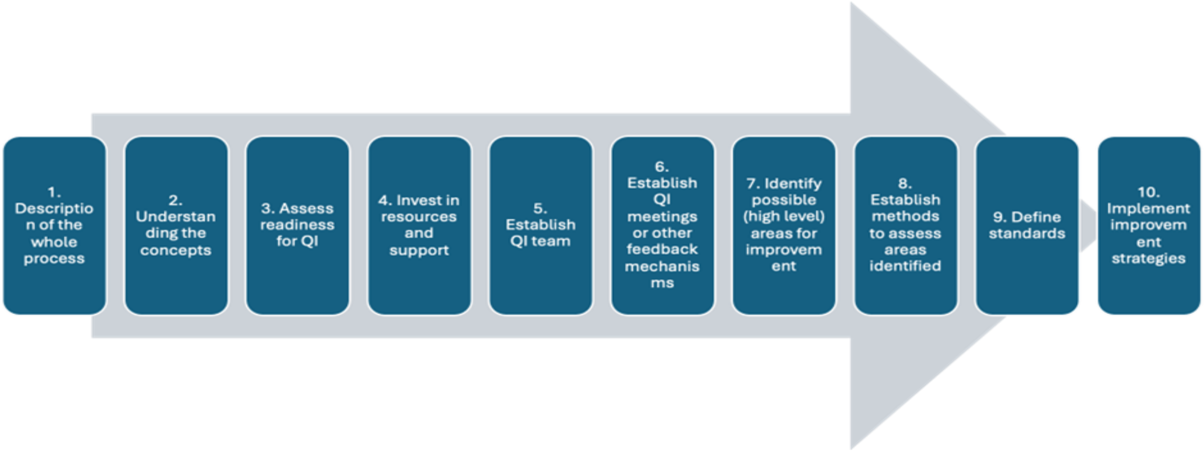

*Figure 2.1 - Linearized conceptualisation of the QI process consisting of 10 stages*

This was used as a framework to identify existing WHO tools and guidance for QI in the EU by a search of related websites, repositories and databases in April 2024. All findings were included into a matrix (Figure 2.2) which consisted of 10 columns representing the different stages of the linearized conceptualisation of the QI process, while the rows included the 7 WHO quality of care domains and a generic QI domain, which incorporate all general QI related resources.

| | Whole QI process | Understanding the concept of QI | Assessing readiness for QI in facility / ED | Invest in resources / support needed for QI | Establish QI teams | Administrative specifics of establishing QI meetings / Other feedback | Identify possible areas to assess for improvement | Establish methods for assessing areas defined which require QI | Define standards | Implement improvement strategies |
|---|---|---|---|---|---|---|---|---|---|---|
| Generic QI | | | Trauma Registry Checklist. | | | | | | | |
| Clinical Effectiveness | | | | | | | | | | |
| Timely | | | | | | | | | | |
| Safety | | | | | | Focus is on clinical effectiveness, but given process is the same. Tools can apply to other areas. | | Yes, due to some overlap with clinical effectiveness. | Yes, but not in emergency care tools. | |
| Patient centred | Focus is on clinical effectiveness, but given process is the same, tools can apply to other areas. | | | | | | | | Yes, but not in emergency care tools. | |
| Efficient | | | | | | | | | | |
| Equitable | | | | | | | | | | |
| Integrated care | | | | | | | | | | |

*Figure 2.2 - Matrix on the resources available for QI for the WHO QI domains.* *– No resources available.*
*– Domain specific resources available.* *- Resources available but not specific to domains.*

#### Supplementary Materials 03

##### Findings from qualitative interviews – implementation of QI activities in the Emergency Units - Phase 1 of the study.

*Implementation and support for QI initiatives*

There was a QI champion in 6 EUs, while 10 EUs had dedicated teams focusing on QI; seven hospitals had hospital-wide QI teams whereas in one hospital, there was only one focal person with responsibility for overseeing all QI activities including those of EU (Figure 3.1). EU QI teams typically included emergency physicians and ED nurses, with involvement from Information Technology (IT) support, administrative staff, and other relevant personnel depending on the initiative's focus.

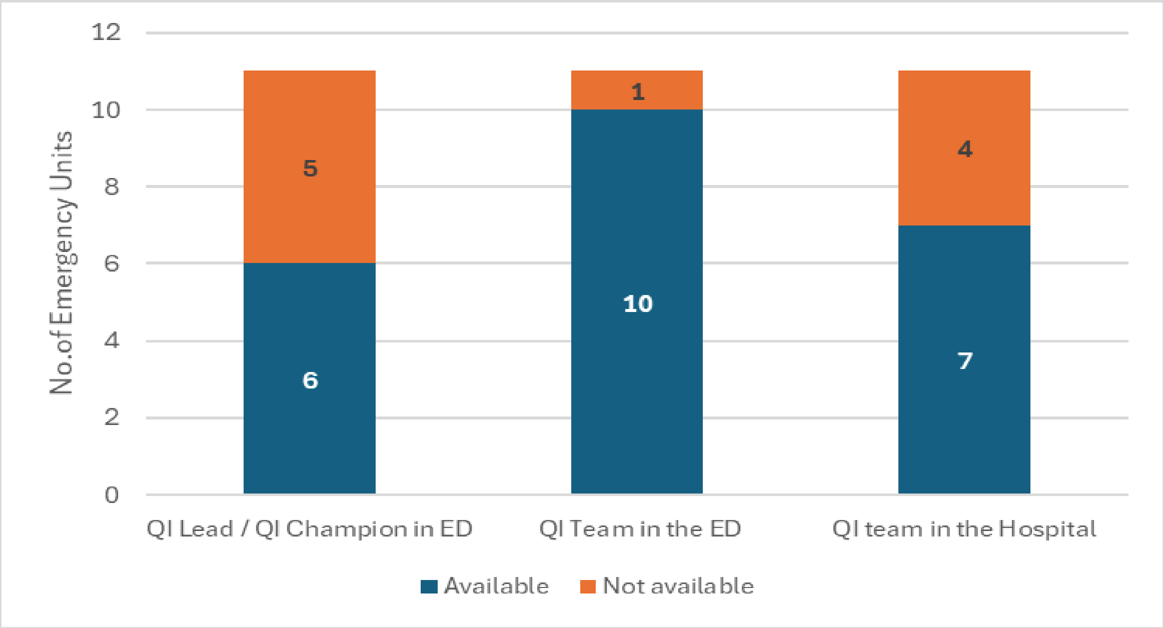

*Figure 3.1 - Availability of QI Leads / Teams (N=11)*

Of the 10 EUs with a dedicated QI team, 9 held frequent (quarterly [2], monthly [5], or weekly [2]), meetings, while 1 had meetings only occasionally when there was a perceived need for a discussion. The support from EU staff to conduct QI activities varied, influenced by workload and availability of staff. Of the 10 respondents who discussed support from other departments in the facility, the majority (6) felt there was some resistance for the EU QI activities (Figure 3.2), while most agreed their management was generally supportive of the ongoing QI activities in their EUs.

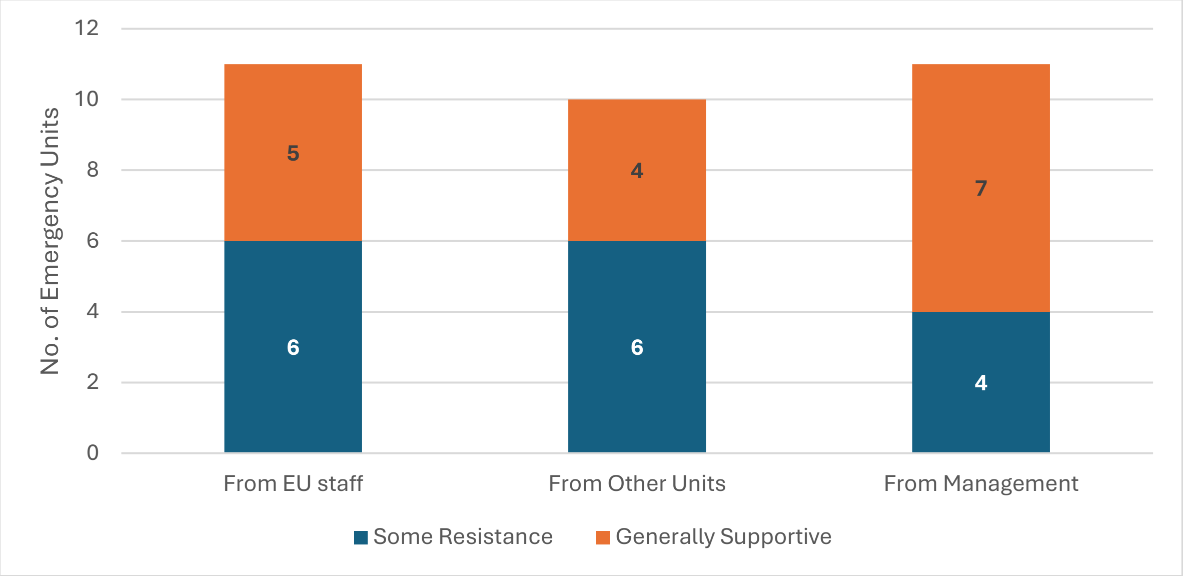

*Figure 3.2 - Perceived support / resistance for QI activities from the EU, from other units and hospital management (N=11 but only 10 responded for support from other units)*

##### *Evaluation and improving quality of care in EDs*

Several approaches to evaluating quality of care were described (Figure 3.3) which focussed on QI initiatives to address effectiveness, safety, timeliness of care and patient centeredness. These included reviews of mortality figures at regular morbidity and mortality meetings, focussing on preventable deaths; monitoring key performance indicators (KPIs) such as patient flow and response times; and use of formal processes for reporting and analysing patient safety incidents and near misses. Concerns regarding infrastructure issues and staff performance related to patient care quality were discussed with a view to initiating corrective actions (e.g.: informing management or additional training and protocol reinforcement for staff). Although patient feedback was not consistently sought, respondents acknowledged its importance in delivering quality care, while some indicated their attempts at QI based on patient complains, though no formal response processes exist. External reviews were also utilized for unbiased evaluations as needed.

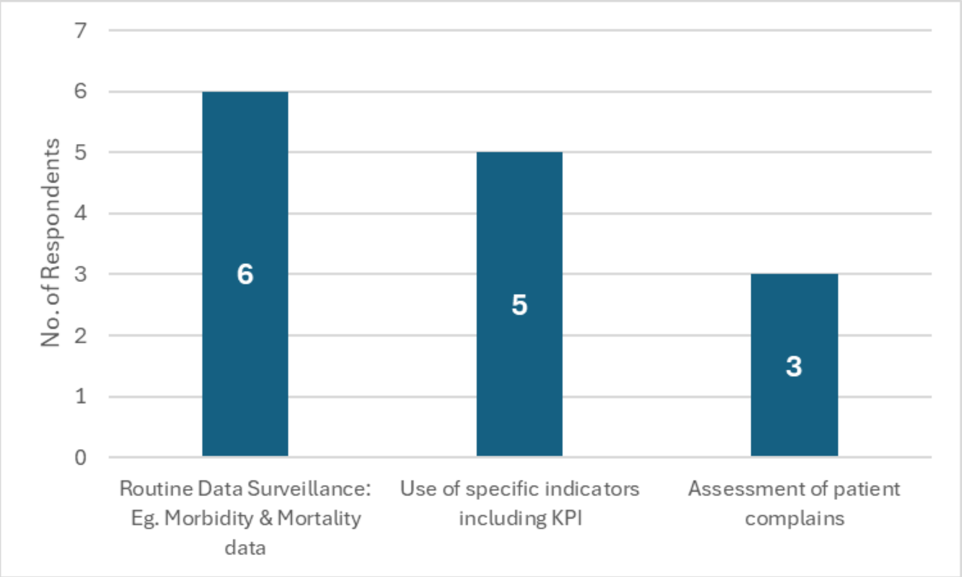

*Figure 3.3 - Common methods used for Monitoring of QI activities (N=11)*

While all EUs utilized some form of data collection for QI purposes, only four specifically used trauma registries. These were tailored to individual hospitals or EUs, primarily capturing essential patient data with limited capability for clinical decision-making support. Two EUs were utilizing the WHO Clinical Registry (Trauma Care).

### Supplementary Materials 04

#### Findings on the barriers and facilitators for quality improvement in Emergency Units from the qualitative interviews - Phase 1 of the study.

*Please note - The numbers within the square brackets refer to the relevant illustrative quotes which are listed as a table at the end of the document.*

#### *Section A: Barriers for Implementing QI in EU*

Respondents primarily discussed their perceptions and experiences related to the broader challenges of implementing QI initiatives, rather than focusing on specific efforts. However, when categorising the interview data against the WHO domains of quality, comments were mostly aligned with four key areas: clinical effectiveness, patient safety, patient-centred care, and timeliness. There was no specific information related to equity or efficiency identifiable. Although challenges related to care outside of the EU were mentioned, QI in the domain of integrated care was not specifically mentioned; the discussions were largely centred on the EU setting. Findings are presented below, where possible, within the QI domains, categorised within the WHO Health Systems framework domains. Quote numbers are shown in square brackets and presented in supplementary material Table 1.

##### 1. Clinical effectiveness

###### *1.1. Challenges related to healthcare service delivery*

Numerous challenges were highlighted with regards to enhancing the clinical effectiveness of care in the EUs. Inadequate healthcare infrastructure for implementing QI initiatives, such as the unavailability of essential resources including a stable electricity supply [1,2,3], and the failure of backup power measures (e.g. generators without fuel) [3]. Lack of adequate space and infrastructure, often due to poor planning of the EUs, made it difficult to accommodate the larger patient load leading to congestion [4,5,6,7], further hindering the care quality in the EU.

In some EUs, the presence of old and obsolete equipment [8] and the frequent malfunctioning and breakdown of such equipment further disrupted the implementation of intended QI activities [9]. Poor maintenance of equipment and infrastructure, further contributed to these equipment failures and the delay in repairs [5,6,10].

Frequent changes in protocols both for clinical care and QI affected the consistency of care in the EU. The lack of standard hospital and EU guidelines on the care of patients were a main cause for this, where delays in updating the guidelines, as well as the reliance on different protocols by different clinicians were also a cause for confusion [11,12,13].

##### *1.2. Challenges related to health workforce*

The lack of staff was a major concern limiting improvement of clinical effectiveness, as was lack of necessary knowledge and skills among the available staff [14,15]. Lack of staff recruitment, out migration and staff resignation [16,17,18,19] as well as the lack of efforts by healthcare management to retain health staff were highlighted [18]. Low salaries and poor incentives [20,21], lack of appreciation for the hard work done [22], lack of career progression prospects and opportunities for their own capacity building and development [23, 24,25], were important underlying reasons for staff shortages. Lack of adequately qualified staff led to the remaining staff needing to manage a heavy patient load, leading to exhaustion and further demotivation [26, 27,28,29,30]. These issues sometimes led to resistance toward the implementation of additional QI processes and non-compliance with quality-of-care standards [31, 32,33]. As a result, reducing the high patient load takes priority for staff over delivering high-quality care to patients.

Another important factor highlighted was the frequent rotation of staff within different units of the hospital, where skilled and knowledgeable EU staff are moved out of the EU to other units of the hospitals and new staff shifted into the EU [34]. Fragmented care within the EU was also thought to lead to lack of overall knowledge and skills among the EU staff [35]. In addition to the clinical staff, the lack of skilled support staff, especially for providing infrastructure and maintenance support, significantly impacts the quality of care in the EU [36]. Poor emphasis is given by hospital leadership towards providing adequately skilled staff, mainly due to the existence of a perception that it is more important to provide the needed funds and resources to care for the patient load.

##### *1.3. Challenges related to health information and communication*

Lack of computers and related equipment for electronic data collection and analysis was highlighted [37,38]. Lack of equipment as well as the lack of maintenance of such IT equipment affected high quality data collection processes [38]. An unstable internet connection, low bandwidth and poor connectivity, especially in certain geographical areas was another important factor leading to failure to upgrade IT systems to accommodate electronic-based health records, data collection systems, and databases [39]. Furthermore, lack of compatible software and hardware within the hospital posed a challenge in utilizing modern data collection software, especially when generating reports [40]. Efforts to address these issues were slow, often resulting in them being forgotten over time [41]. A key factor contributing to the IT and connectivity problems in state hospitals was the lack of well-trained and knowledgeable IT support staff. Hospitals' inability to offer competitive salaries and attract skilled IT personnel hinders IT-related QI initiatives both in the EUs and throughout the hospital [30, 42], and as a result, many hospitals continued to rely on the inefficient paper-based hospital data collection systems [43]. Since data collection is usually done by the EU clinical staff, these inefficient data collection methods become time consuming, and when combined with staff shortages, makes accurate data collection increasingly difficult [44].

##### *1.4. Challenges related to availability of essential medications, vaccines and health technologies*

Poor availability of required essential medication and consumables, mainly due to lack of dedicated funding, was another important challenge [45, 46, 47]. So was the lack of essential diagnostic equipment such as CT scanners [48], which posed a significant challenge to patient care quality, with patients requiring to be transferred to far away hospitals, for diagnosis.

##### *1.5. Challenges related to funding*

A major underlying challenge for QI is the lack of adequate funding for implementing QI activities, which is also the basis for several other reasons related to challenges for QI. The primary issue for this situation is that these hospitals are heavily reliant on government funding, with minimal funds being generated by other means [49, 50,51]. The government funding allocations for public hospitals are barely adequate for the routine functioning of the hospitals and towards covering the recurrent expenses, leaving no funds for QI activities [52,53]. In most cases, any remaining funds get allocated to clinical units for activities to provide greater patient care rather than improve quality of care. Occasionally, even the funds which are initially allocated for the EUs are redistributed to other departments, depending on the intended priorities of the hospital management [54, 55,56]. Thus, securing funds for QI activities in EUs was a significant challenge in most hospitals, often requiring repeated proposals from EU leads to request financial support [57,58].

##### *1.6. Challenges related to healthcare governance*

In general, it was thought that all intended QI activities should be communicated to the hospital management, and their approval obtained. This posed a challenge as hospital management often prioritized serving to patient numbers over improving care quality [54,55]. While management may support QI activities that did not require extra resources, the respondents believed management became resistant when additional funding or long-term investments were requested [59]. This resistance was also perceived to be due to poor training and knowledge about QI amongst healthcare management personnel [60,61], impeding their decision-making in this regard [61]. The issue was thought to worsen when clinicians, rather than trained managers, are asked to take on hospital leadership roles. Additionally, frequent management changes [62] and external influences from the health ministry or regional health authorities [63, 64] further disrupt QI activities at both the EU and hospital levels.

Management perceptions of QI affect the rest of the hospital and health care staff. Frequently, when the EU tries to implement QI activities, other units which need to work in collaboration with the EU will also resist, such as not taking over patients or not continuing with QI processes implemented in EU [65, 66]. Although certain underlying factors, such as staff shortages and lack of adequate facilities within these other units can also lead to such resistance [67], some respondents also felt that a more important reason was the blame culture which persisted within some hospital units [68]. The situation is sometimes so severe that open criticism and negativity are directed at units striving to improve care, with patient complaints sometimes being used to attack other clinical units [69]. This negative attitudes of clinical leads and hospital management contribute to fostering a harmful culture, hindering efforts to enhance clinical effectiveness in patient care.

#### 2. Patient safety

Many of the challenges mentioned above under clinical effectiveness, also apply towards attempts at improving other domains of care quality as well, such as patient safety, timeliness etc. Therefore, the emphasis is given below to certain domain-specific challenges that were highlighted under these other QI domains.

##### *2.1. Challenges related to healthcare service delivery and access to essential medicines*

The respondents highlighted several challenges, particularly lack of patient safety and risk identification guidelines as well as processes for staff to report safety concerns. An important problem was the lack of availability or functioning including basic safe patient stretchers [70]. Lack of proper mechanisms for reporting adverse drug reactions was also reported as a challenge. A frequent challenge encountered in most settings was the lack of a safety culture among the healthcare staff in the ED and across the whole hospital [71]. This was reported as being more prominent in rural hospitals, where the staff have less exposure to new knowledge and developments [72]. Furthermore, the lack of essential medicines in the crash carts, along with the poor maintenance of crash carts could pose a significant threat to the life of critical patients [73, 74].

##### *2.2. Challenges related to health workforce*

The lack of investment in knowledge and skills was identified as an important problem [75,76]. The reasons underlying this were similar to those mentioned above, such as the lack of opportunities for capacity development and skills enhancement, as well as having demotivated staff who resort to easy but unsafe practices in patient care. Poor attitudes among senior doctors and nurses in the EU also contribute, as they often assume superior knowledge and disregard safety protocols [77]. Resistance to improving quality and patient safety was noted by several respondents, largely due to the excessive workload on understaffed teams. Additionally, the absence of qualified QI leads in the EU and hospital was highlighted, leaving clinicians with limited QI knowledge to implement improvements out of a desire to enhance care quality in their units [78,79,80].

##### *2.3. Challenges related to information and communication*

Some respondents also identified the absence of formal patient safety incident reporting systems, lack of guidelines and protocols for patient safety in EUs and the absence of tools and monitoring systems for monitoring patient safety as important challenges [81]. Some hospitals and EUs lack adequate systems to mitigate potential safety risks.

##### *2.4. Challenges related to funding and healthcare governance*

Challenges in securing financial support for patient safety activities mirrored those encountered in improving clinical effectiveness, largely around lack of funding and financial backing for initiatives. A significant hurdle was the management's decision-making on resource allocation which affected the implementation of patient safety programmes. As explained above, poor managerial perceptions on the importance of QI made it difficult to advocate for patient safety activities and secure management support. Often, a serious patient safety incident would have to occur before the management recognized the need to address patient safety risks [70,82]. This issue was compounded by the lack of management involvement in hospital quality and safety committees. Additionally, the absence of accountability frameworks further undermined adherence to patient care standards, with both the hospital management and some staff lacking responsibility and accountability in maintaining standards [83, 84].

#### 3. Patient-centred care

##### *3.1. Challenges related to healthcare service delivery*

Respondents highlighted several challenges for QI initiatives, particularly in integrating patient preferences and feedback into clinical decision-making and care delivery. Capturing patient complaints was challenging due to the lack of effective feedback systems [85], knowledge in how to analyse and difficulty in developing indicators to quantify complaints, as well as lack of knowledge of how to address complaints related to patient centred care. There were also perceptions of a lack of knowledge amongst patients about the care they should receive [86]. These issues culminated in difficulty justifying QI programmes to enhance the provision of patient centred care.  Furthermore, even if patient feedback and complaints were available, the absence of effective mechanisms to incorporate such feedback into decision-making and healthcare processes, made it challenging to balance clinically focused improvements with patient-centred approaches [87]. In most hospitals, the focus in the EU is on addressing patients' issues as quickly as possible, rather than providing patient-centred care. It was felt that tools and guidance to enable patient-centred QI initiatives would be of high importance, especially for the advocating of QI programme implementation in EU.

##### *3.2. Challenges related to health workforce*

High patient load, staff shortages, and limited emphasis on interpersonal skills make implementing patient-centred care difficult [7]. Overworked and demotivated, staff often focus on completing tasks rather than accommodating patient preferences [88]. Additionally, insufficient training and focus on patient-centred care further hinder its execution [89]. Respondents noted that without proper education, healthcare providers struggle to prioritize and effectively implement patient-centred care practices.

#### 4. Timeliness of care

##### *4.1. Challenges related to service delivery and health workforce*

A common issue related to delivery of timely services per se was the difficulty in implementing timely operational processes such as reducing the time for patients to see a clinician, mainly due to the high patient loads and limited numbers of EU clinical staff. This leads to queues, delays in being seen by a clinician and challenging to maintain efficient and prompt service delivery [90, 91].

##### *4.2. Challenges related to health information*

The lack of accurate and timely data to measure and track time-sensitive clinical actions, such as the time to administer antibiotics for sepsis or the time to perform an ECG, were identified as major challenges [92]. The respondents emphasized that without reliable data, it was difficult to monitor and improve the timeliness of care effectively.

##### *4.3. Challenges related to governance*

The absence of clear standards and regular management oversight for assessing important timeliness indicators hindered the ability to evaluate and improve timely care practices consistently [93, 94]. In addition, failures in policy and governance on inter-unit care of patients cause poor collaborations between clinical units, leading to resistance for taking over patients or for shared management, thereby denying timely care for EU patients [95]. These governance challenges further exacerbated the difficulties in ensuring that timely care was provided to patients.

#### *Section B: Facilitators for implementing QI in EU*

The study respondents highlighted certain facilitators for QI which they have encountered in their hospitals and EUs. These were expressed for delivering any QI in EUs, rather than towards a specific quality domain. Below the findings are categorised per WHO Health Systems framework (Table 2)

##### *B.1. Facilitators for QI related to healthcare service delivery*

It was mentioned that on most occasions, the national or regional level health administration such as the Ministry of Health or Regional health authorities, have standards of maintenance and care for infrastructure and equipment. These can be requested and used as monitoring indicators for performance and maintenance of the infrastructure and equipment in the unit [FQ1] and will be a useful tool for QI activities pertaining to infrastructure and equipment.

A suggestion to overcome the barrier of implementing improvement strategies due to lack of equipment was to utilize alternative resources. An example was an EU with no X-ray availability where the staff were trained to use the Ultrasound Scanner machine for some diagnoses [FQ2]. This had reduced the patient waiting time. Another participant's perception was that several small, low-cost QI initiatives can be included together to create a bigger QI project, and which address several domains of quality of care at the same time. This might be more effective in getting better outcomes of the QI process than focussing on serial implementation of QI projects [FQ3].

*B.2. Facilitators for QI related to health workforce*

Enhancing the knowledge and skills of the EU staff was thought to be an easy but very important QI activity, which can be implemented within the EU without requiring any additional funding. Some EUs had their own internal training activities [FQ4], while some arranged even external training for their staff [FQ5]. Suggested were monthly knowledge sharing sessions or practical skills training sessions, similar to Continuous Medical Education activities; in addition to increased knowledge and skills, it was thought that these activities would strengthen the team mentality of the staff, motivating them to work towards achieving better QI care. Another important input was to integrate training into a monthly QI meeting and with all staff of the EU; it was also suggested that at these meetings internal conflicts could be openly discussed by the EU leads and solutions provided [FQ6]. Additionally, acknowledging staff who have worked hard towards enhancing the quality of care in the EU, during these unit meetings was suggested.

*B.3. Facilitators for QI related to Health information and communication*

Communication among EU staff is an essential component to ensure transparency of the intended activities with the staff [FQ7]. Furthermore, it was thought to foster ownership of the QI activities in EUs, rather than these being seen as led (owned) by the QI or EU leads. Ensuring that QI activities within the unit have clear instructions and well-articulated protocols acknowledged by all relevant staff, will reduce ambiguity and minimize conflicts and resistance to the planned QI activities [FQ8].

Other suggestions around improving communication were for improving care quality per se, rather than delivering quality improvement and it wasn't clear in some cases whether the care-quality improvement strategies was introduced as part of a quality improvement programme. Modern communication methods can be effectively used for to improve quality of care delivered in the EUs. Some facilities had started using telemedicine to address the shortage of specialists, where one emergency medicine specialist can overlook and advice the doctors of several EUs, without being necessarily present at the site of care [FQ9]. Another method was the use of social media and texting services to form small groups among the EU staff, for quick messaging and updating of patient conditions etc., [FQ10] which can improve the communication between staff, even if they do not meet in person.

##### *B.4. Facilitators for QI related to Health Financing*

To overcome barriers around lack of finances for QI, respondents suggested activities which would enhance quality of care with the minimal need for extra funding, such as implementing projects which are small-scaled and incurring minimal cost, rather than try to initiate large scale QI projects [FQ11]. The impact from such small initiatives can be quite significant in improving the quality of care in the EU. Activities such as introducing a simple patient triage process for EU admissions can reduce patient queues and enhance patient centred care, as well as timeliness [FQ12] at almost no cost, while organizing monthly CME programmes for the EU staff can have a high impact on clinical effectiveness of care, with no requirements for funding [FQ4].

Towards seeking additional funding for QI projects, the staff can utilize their abilities and connections to access relevant Non-Governmental Organizations (NGOs), private sector or setup collaborations with other larger hospitals and universities at national level or international level which have funding capabilities, to plan and initiate larger QI projects [FQ13].

##### *B.5. Facilitators for QI related to healthcare governance*

It was thought that effective communication and advocacy were needed to ensure management consent for QI activities, especially for implementing low-cost initiatives [FQ14]. Furthermore, when the management sees the positive feedback from patients and the public for the QI initiatives, they often become motivated to support such projects over others [FQ15]. Towards reducing the challenges posed for QI activities from other units, development of protocols or guidelines together with other relevant unit heads, specialists, and hospital management on owners of processes was thought to be beneficial. In this way, most of the inter-unit conflicts can be minimised and the quality of patient care can be improved [FQ16].

#### *Section C: List of Quotes*

*Table 4.1 - List of Participant Quotes on Barriers for Implementing QI in EU*

| Quote No. | Interview No. | Quote |
|---|---|---|
| 1 | IDI-2 | We don't have a stable electrical supply. So, for 2 hrs at a stretch 3 to 4 times a day, sometimes we don't have electricity, and when that happens, we have to switch on to generators. |
| 2 | IDI-6 | …basic necessities like electricity, or running water, is a luxury here. |
| 3 | IDI-7 | But when the national grid electricity is off, we usually get a backup generator, but the backup generator in many cases do not have fuel all the time. |
| 4 | IDI-3 | …there are patients who are then kept in the emergency department after admission and that's what we call boarding, and boarding is an issue in our ED. That, I think, is the main challenge really, and we don't know who to blame. |
| 5 | IDI-9 | …a lot of infrastructure weaknesses, because it wasn't built in from the beginning, of how to maintain the infrastructure that was built and who's going to do it? |
| 6 | IDI-9 | Unfortunately, because a lot of these hospitals' maintenance plans are not put very well. And today's there's a lot of support, and it might be a good fund to build a very nice, shiny new hospital with a lot of nice fancy equipment. But then there's absolutely no plan for maintenance. |
| 7 | IDI-3 | Capacity is the main challenge from our side. There are more patients than we can handle, or the hospital can handle. |
| 8 | IDI-11 | …the infrastructure itself is sometimes very basic or non-existent. So even if you have the personnel, I mean they wouldn't. They wouldn't serve in supporting you, because there's nothing to work with or to use. |
| 9 | IDI-9 | …these amazing infrastructure that was there, but no one is there to maintain it. No one is there to oversee it. There's no long-term plan for it. So over time it dials out, and then no one actually ends up knowing what you have in hospital. |
| 10 | IDI-2 | …CT Scanner is needed to improve quality because it's broken, or whatever, that won't come from the hospital, so we have to wait them for a national procurement plan which comes from province. |

| | | |
|---|---|---|
| 11 | IDI-5 | First the QI, and the treatment method. It changes very fast. Right now, we do a lot of research, and another QI keep coming up, come up and sometimes we don't know if it is the right thing, it will come this year. In another 2 years, they say no, we don't do this anymore. It changes very fast. |
| 12 | IDI-9 | …debates around what procedures should be done by who. So, there's a lot of cross departmental sensitivities a lot of the time with these primary teams. That approach was developing a shared protocol that all of the heads of departments would look out, and all of the heads of departments would sign up on this protocol. and then it would be propagated to their different departments. so that there is a there's a document to resolve most of the day-to-day issues. |
| 13 | IDI-1 | If you have a leadership that is running an emergency department that is fragmented, with multiple entry points, where the ER, you have to go through that way, then another emergency department has got 4 areas. And then an emergency department has got to 4 leaders of different specialties, and the other person does something else. And then that's fragmentation. It's very difficult to lead the people who are working in the front-line because you are going to have different leadership streams. |
| 14 | IDI-1 | We had human resources skill issues. We had human resource numbers issues, and this has still been a challenge. |
| 15 | IDI-1 | …so that awareness raising, that awareness would be the main challenge in lower- and middle-income countries. |
| 16 | IDI-6 | …this as Africa, there will always be brain drain. People will take exams and travel and leave Africa because you can be called a doctor, but you are unable to pay your child's school fees, for example, because you've been paid so little. |
| 17 | IDI-9 | …government salaries are very, very, very low. So, this person isn't really motivated. There's not a lot of accountability framework on his production cap on what he actually does or if he doesn't. So, there's accountability in him coming to work, for example. attending and leaving at the end of the day but what he does in the 8 or 9 hours of work, not a lot of people would keep an eye on that. And then, and he's not incentivized to create change both at a personal level, because it's not getting paid well enough… |
| 18 | IDI-1 | Because a lot of specialists are trained, but retention has been the weakest link. We have specialists that have been trained by the government. But retention of these specialists has been the weakest link. Some of them come back but the emoluments are paid, maybe for junior doctors, and in that way, it makes them seek greener pastures. |
| 19 | IDI-5 | …the nurse turnover is high, because mostly they come like a 2 or 3 year to practise their skill. After that they go to the private hospital because the work from the government hospital will qualify as experience… |
| 20 | IDI-6 | We are losing doctors in this area. The lucrative incentives for public health doctors, public health specialists, in [this country] is at least 5 times what a regular doctor’s making. |
| 21 | IDI-9 | So, the staff, we have a very big problem of turnover. In these regions there's it's low pay, and most people are only staying long enough to get enough credentials, so they are able to leave the country or leave the government sector to be able to work in a better-paying private job. So, a lot of the time there's unfortunately not a lot of retention, and that creates this gap. |

| | | |
|---|---|---|
| 22 | IDI-9 | …if there's no quality, it's done only because the staff would like to see it being done. But there is no reward of it now being done, and there's no punishment if it wasn't done. So, that doesn't help incentivize hospitals. |
| 23 | IDI-11 | I would expect that in some settings also, the capacity in terms of the abilities like the personnel, level of training and education regarding quality and quality improvement can be challenging. |
| 24 | IDI-9 | …eventually you find them to be burnt out. There's no satisfaction in the work they're doing. They don't see it going anywhere. They're very depressed, and they feel like. Well, it's, I just have to come to work and leave at the end of the day. They don't really want to bring out any change, even if they didn't start that way. So, it's very unfortunate in that end in that way. And they did. And they say there's a dead end. There's no career progression for them. There's no ability for them to technically improve in their field. There's no continuous training. So, all of that adds to the mix as well. |
| 25 | IDI-2 | We don't have regular capacity programme for the now. |
| 26 | IDI-11 | …in the lower resourced settings where there's not so much capacity, there’s not a very well-structured team to be responsible for quality improvement. |
| 27 | IDI-2 | We have sporadic visitations from the quality department. But then, we had a recent staff shortage, and so, people from that department even moved. And so, there isn't the time or capacity to even do something like this, which is important. |
| 28 | IDI-9 | The quality improvement team in the hospital, a lot of the time are not conditioned. And they don't really have a lot of clinical background. And in my region of the world, they're often not very strong technically, because it's not a priority that the hospital would deem. |
| 29 | IDI-9 | …also, one of the biggest challenges is, there's not a full-time dedicated quality leads. So, it's more the additive responsibilities to variably basic clinicians already, for them too, but it is only because it was only successful relatively because they wanted to see change. |
| 30 | IDI-9 | People in the hospital and I realized they've got very, very strong infrastructure, but none of it is in place. But it starts somewhere, and it was very simple costs. But the hospital management didn’t want to pay that cost because they're like, this is like the last of our problems is to support the IT. |
| 31 | IDI-11 | There are some key issues that whenever you would try to implement in a low resource setting in some of the places, you would find some resistance, because they feel that this is so much lower, it's this is so much work for them, and that they cannot like, keep on doing this. |
| 32 | IDI-10 | …if you don't remind everyone then people take it very easy. They start ignoring things that happens. That's not related to new staff or anything. It's just the old staff losing their motivation. |
| 33 | IDI-11 | They're more resistant and sometimes the resistance, because they think that this is not a priority, or that they don't have time, or that they have, they are already overworked, and sometimes because they just not convinced that this is a priority or something that's with me. |
| 34 | IDI-9 | …a lot of these departments would have rotating staff. So, the staff would come in for like a period of 2, 3 months, and then they would completely be replaced, and new staff comes in, and by the time they get settled, and a lot of the time, there's not a lot of time for let's say initiation for them. So, they're not properly briefed before they start working. |

| | | |
|---|---|---|
| 35 | IDI-1 | …but the care is still fragmented. Why? It's because the human resource skill is lacking. |
| 36 | IDI-8 | The challenge is really more about the resources that are available. So, having said that it's not mainly about, perhaps finances or money, but it's also about capacitation of competencies. |
| 37 | IDI-6 | …lack of computers and electricity in most of the facilities… |
| 38 | IDI-4 | …there's always an internet issue and the tablets that they collect the data with, there's a problem with the, you know, utilization and maintenance and all. |
| 39 | IDI-7 | And then also the issues of the internet. And so, our EMR is in the local area network. So even if there is no internet, it can continue to work. However, when the internet is connected, usually the bandwidth is small. So, for the process of data entry takes like, it's much, much slower… |
| 40 | IDI-7 | …the hospital acquired an electronic medical record. Yes. Now the problem with that is that we cannot extract information we need for our clinical and mortality audits from that specific EMR. |
| 41 | IDI-7 | …the people who provided the electronic medical records to make sure this form is part of their system. Yeah. And that takes a very long time, because I talked to them way in February. So up to now and yeah, never done anything. |
| 42 | IDI-9 | …even if the hospital had initially procured a lot of very nice laptops and computers and batteries and UPS systems and servers and internet connections. The person that would be responsible for that is someone that is not very well trained. It might not be his field, unfortunately, might not be his primary field, because it was never, because the hospital was not being given enough to bring someone that is very competent. |
| 43 | IDI-8 | …currently, there's still a lot of challenges, because some of our data are still collected using paper and pen. |
| 44 | IDI-2 | It impacts on patient care when, in fact, we don't have enough staff for it. So, the idea will be to see where the 2 systems would potentially merge so that it's no extra work for the people putting things on the floor. |
| 45 | IDI-1 | …we realised that we couldn't even afford a basic ECG. |
| 46 | IDI-7 | …the biggest concern was availability of drugs and sundries. Yeah. And in that case, because now the budget comes from the central government, we don't really have much to do about it. |
| 47 | IDI-7 | …challenge is to do with the supplies, equipment and drugs. |
| 48 | IDI-6 | We don't have our own CT scan machine. So, what we do: we see these patients, if it is a stroke, for example, or cerebral vascular disease, we stabilise, control blood pressure, and we send for CT with our own ambulance. They are taken, CT is done, then the patient is brought back to us for continued management. |

| | | |
|---|---|---|
| 49 | IDI-11 | …whether you would like to look for funds externally, and there's there is no capacity, or there are not enough people to be willing to work on the project… |
| 50 | IDI-2 | We may have other departments that provide, like NGOs, that provide support for rehabilitation of drug abusers or provide food, but in terms of quality improvements in the hospital itself, we don't have such things. |
| 51 | IDI-4 | …but I wouldn't say much is being done in PPP yet. |
| 52 | IDI-11 | So it could be that everyone is convinced that this is important, but there's not enough fund for it. |
| 53 | IDI-2 | …we have major financial limitations. Just as we're speaking now, we're receiving information from national treasury that we have almost exhausted our budget nationally. |
| 54 | IDI-9 | The government has these vertical budget lines. So, the government has free meditation for emergency care, which means that they will provide certain medications. But if the system isn't strong enough, what ends up happening is, okay, there is a budget the pharmacy team is supposed to procure medications and supplies for the month but if the if the equipment, the supplies they run out. let's say, 2 weeks into the into the department the other 2 weeks. It's patients paying for these medications that the state is providing for free, because again of an administrative failure somewhere a lot of the time. |
| 55 | IDI-9 | …as a senior manager. If you have a lot of money coming into one department from your Ministry of Health, and you've got other problems in other departments that generate income for you. It is very hard to make that argument. but in legislation emergency care is free as a total…. |
| 56 | IDI-2 | We haven't been allocated a budget. And so, whenever there's an issue in another part of the hospital, whatever is not deemed absolutely necessary, funding will be moved there to transport it out. |
| 57 | IDI-6 | …need to rewrite the proposal and send it again to get more funding. |
| 58 | IDI-2 | We have to actively seek it and come up with once again a proposal, and then we've got a hospital barrier to get through, because we have to prove that it can't come from our hospital. We don't have great working relationships with multiple Prehospital providers, including private paramedics. So, there's no QI Forum. |
| 59 | IDI-11 | They [management] are usually supportive. But it depends on what we mean by support. I mean, they are usually for the project. They would like it to be (…) implemented, but sometimes, when there are some challenges that are very difficult to address or that are not a priority from their perspective or point of view, (…) they may not like be actually actively supportive |
| 60 | IDI-9 | …it doesn't help when the lead of the hospital isn't very technically proficient at that, and the hospital management is unable to hire someone with better proficiencies. |

| | | |
|---|---|---|
| 61 | IDI-9 | In our setting, a lot of the management is not dedicated managers. So, there are people that are clinicians. A full-time surgeon, a full-time intensivist, a full-time in parents is very good at what he does in his clinical, respective field, but then you put him in a management post, a senior management post as a hospital lead or emergency or a unit lead. And then the issue starts to appear because it's not their skill set. It's not what they've been trained to do. |
| 62 | IDI-9 | Another thing in the challenge is our change of leadership. Frequent change of leadership in our settings also creates this. So, change of hospital management or changing of emergency unit management… |
| 63 | IDI-2 | …obviously political, provincial push, has to take preference, because of our sort of service delivery paradigms that we have to see to. |
| 64 | IDI-2 | …we often have a top-down approach where people from Province will come and tell you, this is what you need to do in your system, or this is what you need. These are your outcomes, but they don't work because our system is different, and our factors are different. And we need data on that. |
| 65 | IDI-10 | The support from departments which are outside our control of supervision is quite challenging. It is difficult. We have faced difficulties to implement quality projects because the other department do not believe in the emergency care related quality projects. |
| 66 | IDI-10 | …we do have challenges, and they required meetings, and helping them align to our vision and the patient. So, it takes times. But yes, we have challenges regularly when it concerns other specialties. |
| 67 | IDI-3 | Sometimes you get, you know, pushback from some other departments, because they might not have resources, or their processes might not align with the processes in the emergency department. |
| 68 | IDI-2 | …without proper management oversight, when we have combined meetings that make the sort of mortality, there runs the big risk of passive aggressiveness or defensive culture, where, instead of addressing the complaint in a psychologically safe manner, often times it becomes one department against another which becomes a barrier to good quality improvements. So, it distracts from the actual message of what's going on, |
| 69 | IDI-2 | …difficulty we have is the understanding of complaints and the importance of reporting them because, there still exists this blame culture or perceived blame culture which prohibits people from actually reporting things. And this is not just our department. This is from the top down, from everywhere. It's perceived that the more complaints you have, the worse your department functions, when, in fact, it's the opposite. |
| 70 | IDI-2 | …patients fall from the bed because the stretcher doesn't work, |
| 71 | IDI-2 | Like currently when things were wrong, it's not really a surprise, but people don't do anything to stop it… |
| 72 | IDI-1 | …in the rural [country name] hospital, is that it's first of all to understand that most of the things are not practiced the way I would love them to be practiced. Because this is a rural province, and, somehow, you need to be a little bit patient, and to understand that people have not had the exposure you are talking about. |
| 73 | IDI-1 | This was also a big area where crash carts were always empty. They were always empty. They didn't have a system of managing a crash cart to replenish what was lacking. |

| | | |
|---|---|---|
| 74 | IDI-4 | …so, availing the emergency essential drugs and equipment. So, it touches everything basically. |
| 75 | IDI-1 | You can have a structure. but you can't make use of it and miss a lot of diagnoses and have more problems because you have not invested in a skill set. Why don't we have a skill set? This is usually a policy level. |
| 76 | IDI-7 | It is very common in our environment to have equipment, actually that you don't even know how to use. Same with a structure. You can have a structure. but you can't make use of it and miss a lot of diagnoses and have more problems because you have not invested in a skill set. |
| 77 | IDI-10 | We have attitude issues, especially from senior staff. so, they may not agree with certain, I mean, they may deviate from the protocol and feel like it's their right to deviate from the protocol. So, this tends to happen with very senior specialists or very senior nurses. |
| 78 | IDI-12 | The level of resistance is different from one setting to the other. So, when we're talking about a setting where, like the daily ER visits is very high and patient flow at some, especially at some points of the day, or like in during the very highly high, high flow hours of the days you would find more resistance than in other settings. |
| 79 | IDI-11 | It would just need some more communication, and like discussion to keep them on board. And it's sometimes more difficult in these settings than the others. Just to get everyone on board. You would get some at the at the end, and some would stay resistant, and maybe like join later. |
| 80 | IDI-11 | There has always been some resistance to improve the documentation practice in some settings. well, actually, I would say that there was there. There's been some resistance in improvement in initiatives to improve documentation in all of the settings. |
| 81 | IDI-2 | …trying to improve the reporting structure, because it's quite irritating the way it's done, where you must go and find a form and do this and do this…. |
| 82 | IDI-2 | …and obviously we get far less support if an incident hasn't yet occurred. So, to prove why this is needed takes a lot more effort, because there's multiple other competing interests in the hospital, given where we are, with resource limitations. |
| 83 | IDI-9 | So, it creates a lot of wastage where you've got amazing infrastructure, but no one there to really maintain it, oversee it. And again, no accountability framework whatsoever. |
| 84 | IDI-9 | …weak governance and lack of accountability, frameworks also create a big issue around that, and to be honest is also the element of lack of incentivization or quality improvement. |
| 85 | IDI-1 | I'm also very interested in complaints. Although these are very difficult to measure from our side as end users and front-line doctors, we rely on what managers are receiving in the morning brief… |
| 86 | IDI-10 | See in our place, patients are quite ignorant about quality systems and quality process. It's very rare that a patient would be educated enough to understand quality systems. So, but we do let them know that they are under a certain pathway, |

| | | |
|---|---|---|
| 87 | IDI-8 | It's also about a bottom-up approach, perhaps, and really listening to those who are working in the EDs and in the EUs, and what they have to say. And of course, the centre of it all is patients. So, we also have to listen to what the patients may have to say in this. Our thoughts, our perspectives are mainly clinician focused, we may have yet still to hear from the patients, at least in this side of the world. |
| 88 | IDI-7 | This staff were a bit, and not so kind to them. Those ones, we do have sessions on customer care… |
| 89 | IDI-1 | We had to sit to share the benefits of triage and to introduce them to what triage is, because it is not something that they were familiar with. It is something that was new to them. |
| 90 | IDI-1 | …though it is meant to be a tertiary hospital and a referral hospital, it is looking after everything, including flu, you know, including common cold. And there were so many patients in the emergency department and a lot of complaints of delays in being attended to. |
| 91 | IDI-3 | There are others who might find it burdensome. They might find like, we are the only one who's being asked, and other people are not playing the balls. They feel like they are being the sort of targeted or singled out when other people are not willing to make the change. |
| 92 | IDI-1 | …one of the quality things we would want in an emergency is how quickly we attended to very sick patients. Meaning those triaged, you know, red, orange. We want to look after them very quickly, and how quickly you attend to them becomes a very important thing for us. Because we don't want someone who is triaged red, acutely ill, to be seen 2 hours later, 3 hours later. I think, then that means we are failing. |
| 93 | IDI-2 | So, I'm putting a lot of my effort in there to make data driven decisions. So, we haven't had that data available before. Now, if we design the system with QI and data driven decisions in mind, we should be able to collect the right amount of data to inform staffing decisions, resource allocations, the number of near misses and using metrics and early warning things to pick up from the data to see what we can do to inform actual QI plans. |
| 94 | IDI-8 | As for the metrics, it's the same story. Different facilities may have their own analysis in their own different ways, as to their turnaround times, even volume of patients, metrics, and everything. But not on the national level. |
| 95 | IDI-11 | …I mean, you may find also, if we're talking about improving the length of stay, or improving like the time to consultation or so, sometimes you would find resistance from the others, apart from the other specialty… |

*Table 4.2 - List of Participant Quotes on Facilitators for Implementing QI in EU*

| Quote No. | Interview No. | Quote |
|---|---|---|
| FQ1 | IDI-5 | For the structure, we actually have some standard and some self-assessment tool that the Ministry of Public Health sent to us, and we have to send them the information on what we have. |
| FQ2 | IDI-2 | …we have an issue with 24 h radiology services. So, one such programme was to try and supplement by improving ultrasound use and utilization for certain conditions that we don't have access to X-rays for… |
| FQ3 | IDI-4 | We have one project which works on all of this, like to improve the emergency care systems. So, it basically includes to improve the triaging system of the emergency department and the capacity of all health professionals, to train all so emergency care providers on the basic emergency care, to dedicate an emergency resuscitation area in the emergency department, also availing the emergency essential drugs and equipment. So, it touches everything basically. |
| FQ4 | IDI-1 | I had to start teaching my team members, meaning my interns, my medical officers, the importance of triage and what the benefits are. And then from there we had to hold departmental meetings, meaning our department, the nursing unit, the doctors, as a department of internal medicine. |
| FQ5 | IDI-5 | …every year, they must go to the academic conference, or some year, they have to rehearse about Advance-Life-Support, Basic-Life-Support every 5 year. So, they have to do this. |
| FQ6 | IDI-1 | …ours is very oriented to the ground. If tomorrow, or the other day I heard that my senior doctor had an altercation with one of the nurses, it is solved in that way, but it forms a topic for our next strengthening meeting. Then we put in conflict management, and like that… |
| FQ7 | IDI-11 | …usually, they would be supportive if it was communicated the right way with them. I mean they, if they were involved from the beginning, and would like felt that as part of the initiative and the team, they would be very supportive. |
| FQ8 | IDI-5 | We share the guidelines, improve our SOP. More than that, in every year, they must go to assess their performance. |
| FQ9 | IDI-1 | …telemedicine is the biggest thing that works for us here as a way of improving how we look after patients. |
| FQ10 | IDI-1 | …we have taken advantage of social media, meaning WhatsApp, online calls, video calls to see how we can improve care of our emergency or acute patients. |
| FQ11 | IDI-11 | I would say that to start depending on the setting, to start small, not necessarily like with huge, big projects that are very, that looks very appealing, and but are actually not feasible to implement. So, to start small, just to introduce the culture so that people embrace the culture of quality improvement. And so gradually they would be more ready to do more for it. |

| | | |
|---|---|---|
| FQ12 | IDI-1 | …and we started teaching our doctors, our nurses, to implement triage. And in that way, we were able now to capture the sickest patients, and we started experiencing less of people dying while waiting to be seen on a queue, or someone collapses on a queue while waiting to be seen. So, we don't have that at the moment. |
| FQ13 | IDI-4 | …we ask the facilities, the health facilities to come up with the matching funds for this project, and also the facilities themselves, try to get some funding from, you know, other collaborating facilities abroad or universities and hospitals… |
| FQ14 | IDI-9 | …very supportive morally and very supportive in every sense that doesn't require a lot of financial commitment. So, because again, it is, it is scarce resources. |
| FQ15 | IDI-2 | …once we have a programme that we believe is community facing and has tangible outcomes, we are well supported. We recently ran a stroke awareness, sort of implementation due to delaying presentations of stroke to the emergency department, and we tried to identify that as an issue and devise the sort of 6-month ongoing programme where we educate communities and try and get feedback and implement programmes and schools and things. So that's still ongoing. So, we are well supported for that because there's a tangible outcome. |
| FQ16 | IDI-9 | So, there's a lot of cross departmental sensitivities a lot of the time with these primary teams. That approach was developing a shared protocol that all of the heads of departments would look out, and all of the heads of departments would sign up on this protocol. and then it would be propagated to their different departments. So that there is a there's a document to resolve most of the day-to-day issues. |

### Supplementary Material 05

#### Findings from the analysis of the eMenti® word clouds on how to measure WHO quality domains – Phase 2 of the study

A thematic analysis of responses is presented in Table 5.1 below.

1. Clinical Effectiveness (38 Responses)

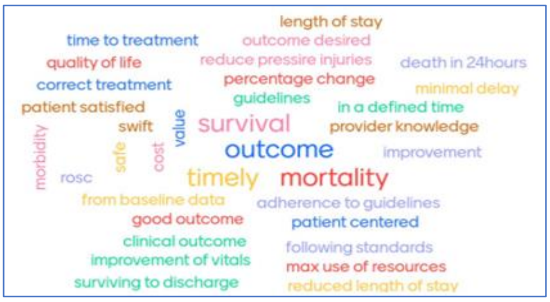

2. Patient Safety (44 Responses)

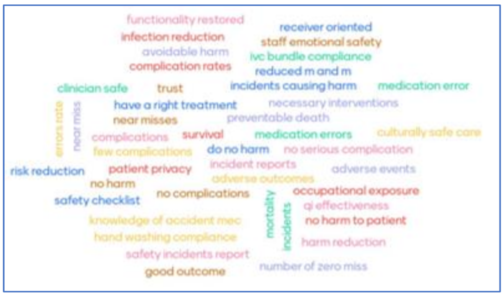

3. Patient Centredness (44 Responses)

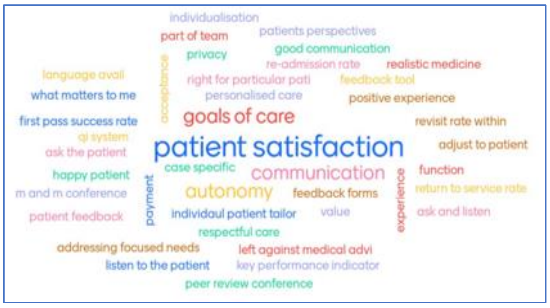

4. Timeliness (41 Responses)

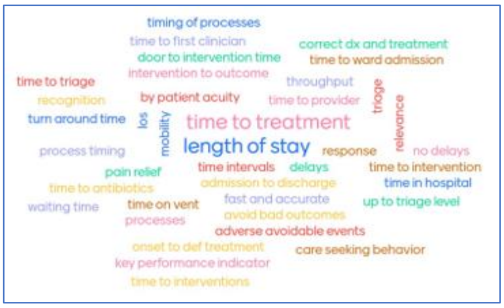

5. Equity (43 Responses)

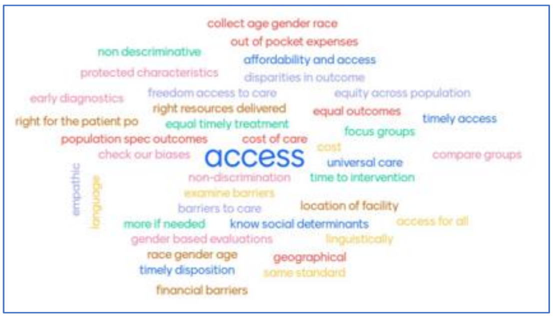

6. Efficiency (49 Responses)

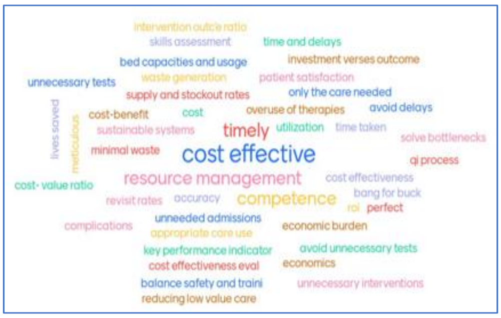

7. Integrated Care (49 Responses)

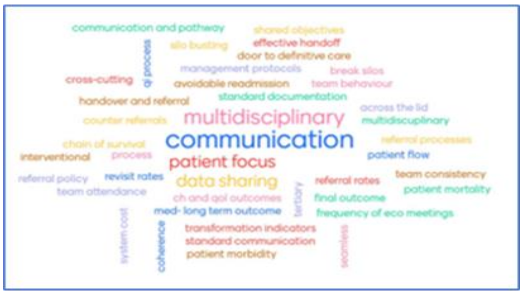

*Table 5.1 - Codes generated from suggested WHO quality domains measures and the numbers of responses*

| WHO Quality Domains | Suggested Measurements | No. of related responses |
|---|---|---|
| **1. Effectiveness** | Clinical outcome | 11 |
| | Timeliness of care | 5 |
| | Adherence to clinical care guidelines | 3 |
| | Mortality | 3 |
| | Survival | 3 |
| | Economics of care | 2 |
| | Length of stay | 2 |
| | Patient centred care | 2 |
| | Clinical Knowledge | 1 |
| | Morbidity | 1 |
| | Quality of Life | 1 |
| | Resources utilization | 1 |
| | Safety in care | 1 |
| **2. Safety** | Safety practise | 16 |
| | Complication metrics | 5 |
| | Clinical outcome | 4 |
| | Errors | 3 |
| | Hazardous events | 3 |
| | Safety reporting | 3 |
| | Adverse outcomes | 2 |
| | Near misses | 2 |
| | Safety knowledge | 2 |
| | Clinical intervention | 1 |
| | Mortality | 1 |
| | Privacy | 1 |
| **3. Patient centeredness** | Patient preference | 10 |
| | Care quality metrics | 9 |
| | Patient satisfaction | 7 |
| | Communication | 5 |
| | Feedback | 3 |
| | Autonomy | 2 |
| | Goals-of-care | 2 |
| | Team meetings | 2 |
| | Case specific | 1 |
| | Experience | 1 |
| | Outcome | 1 |
| | Payments | 1 |
| | Privacy | 1 |
| | Return to service rate | 1 |
| | Teamwork | 1 |
| **4. Equity** | Accessibility | 16 |
| | Non discrimination | 10 |
| | Patient characteristics | 5 |
| | Equality in outcomes | 4 |
| | Timeliness of care | 3 |
| | Correct care | 2 |
| | Empathy | 1 |
| **5. Efficiency** | Cost measurements | 15 |
| | Resources utilisation | 12 |
| | Competence | 7 |
| | Timeliness of care | 6 |
| | Return on investment | 4 |
| | Quality in care | 3 |
| | Patient satisfaction | 1 |
| | Performance indicators | 1 |
| **6. Timeliness** | Timeliness of care | 24 |
| | Length of stay | 5 |
| | Quality of care | 5 |
| | Patient care outcome | 3 |
| | Process time | 2 |
| | Key Performance Indicators (KPI) | 1 |
| **7. Integrated Care** | Integration process | 12 |
| | Communication | 10 |
| | Team effectiveness | 7 |
| | Outcome of care | 6 |
| | Multidisciplinary care | 4 |
| | Process effectiveness | 3 |
| | Policies and protocols | 2 |
| | Patient focussed care | 2 |
| | Indicators | 2 |
| | Cost of care | 1 |

#### Findings from the analysis of the slido® responses on the participants perceptions on the domains most feasible to change – Phase 2 of the study

The table 5.2 below shows the WHO QI domain which the participants mentioned as the most feasible to change (rank 1), in each of the 3 rounds of voting. The second-round voting excluded the first-round rank 1 QI domain, while in the third-round voting, the rank 1 QI domains of both the first and second round voting were excluded.

*Table 5.2 - Number of participants voting each quality domain as the most feasible to change*

| WHO Quality domain | No. of participants responded | | | | Final overall rank |
|---|---|---|---|---|---|
| | Rank 1 in round 1 | Rank 1 in round 2 | Rank 1 in round 3 | Overall approval | |
| Safety | 5 | 5 | 3 | 13 | 1 |
| Effectiveness | 4 | 4 | 1 | 9 | 2 |
| Patient centredness | 2 | 4 | 1 | 7 | 3 |
| Timeliness | 2 | 1 | 4 | 7 | 3 |
| Efficiency | 3 | 1 | 1 | 5 | 4 |
| Integrated | 1 | 0 | 2 | 3 | 5 |
| Equity | 0 | 0 | 2 | 2 | 6 |
| All domains equally feasible | 0 | 0 | 0 | 0 | 7 |

### Supplementary Materials 06

#### Findings on the barriers and facilitators for quality improvement in emergency units from the workshop of technical experts – Phase 2 of the study

Results are structured according to themes identified either because they represented a point in which there was consensus among the group, or they were felt to be important and relevant enough to be considered. Naturally, there is some overlap between themes. Although the discussion was guided to focus on EU specific issues, participants also mentioned general challenges which they thought were particularly prominent in EU settings due to the volume of patients seen and the often-transient natures of the interactions with them, as well as the fact that diagnoses may not have been made during their time in the EU.

##### *A. EU specific barriers*

###### *A.1. Identifying priorities for improvement*

For sustainability, it is essential to adequately identify the priorities for improvement in quality of care; involving staff and service users allows collection of granular data which aids in the process of prioritisation, but this is difficult in the busy EU setting.

- Patient feedback - Asking patients about their experiences of care within the EU, will yield important information about the patient centredness of care quality, if it had been satisfactory or not.
- Feedback from relatives – In addition to the patient feedback, obtaining feedback form the bystanders and relatives of patients who stayed with them would also yield valuable information, especially about the lapses in care quality from the public's perspective.

###### *A.2. Particularities of QI in EU setting*

It was highlighted that unlike the other units of a hospital, the EU setting poses challenges to defining quality of care and understanding how best to improve it.

- Staff safety – staff safety is a particular issue in EU settings which is the first point of entry to the hospital for many incoming patients who may have suffered physical violence. It was pointed out that implementing actions to improve the safety of frontline staff, is a pre-requisite for being able to deliver QI initiatives in the EU.
- Defining of standards of care in the EU, is a particular challenge. A was to focus on improving quality from the current EU baseline, rather than attempting to meet specific standards, although not all participants agreed with this.
- The absence of a diagnosis in the EU makes QI delivery around clinical care pathways difficult.
- Issues identified in the EU might have solutions that lie outside this unit. This requires there to be a broader understanding of the factors which impact upon quality of care in EU.

###### *A.3. Working in resource constrained settings*

Several comments were made on how QI needs to be adapted to a setting where facility resources may not be available to provide the highest quality of care, despite QI efforts.

It was acknowledged that at times, improving the quality of care in the EU in resource constrained settings might require adaptation. For example:

- Adaptation of clinical care protocols to ensure better fit to the EU.
- Attempting periodic data collection or sample data collection where managing a registry or continuous data collection for decision making is challenging.
- Introducing QI concepts to medical trainees and junior staff and encouraging them to undertake small QI activities within the EU, which will improve the overall care quality without a cost nor a staff burden.

##### ***B. Barriers not specific to EU***

###### *B.1. Leadership and governance*

QI based governance and the development of standards, guidelines and regulations, as well as indicators, was highlighted as important for gaining commitment from leadership to focus on improving quality of care. The need for guidance on how to advocate for QI to the hospital leadership was discussed. Although it was considered that clinicians should be the primary advocates for QI, it was noted that their lack of knowledge and skills on advocating might make the attempts ineffective.

Mandating the presence of a dedicated senior management representative as part of the defined QI team of the unit, as well as ensuring that management receive training on QI, were also thought as strategies which could be adopted to gain support of hospital leadership.

*B.3. Challenges in keeping QI simple and do-able*

A common misconception in QI was that it had to be a large and impractical undertaking. Sometimes the desire to make a research project out of a quality improvement initiative, especially with the involvement of academic institutions as partners for QI, led to it being seen as unfeasible. Thus, it was felt there should be particular emphasis on keeping QI simple and realistic to the context in which it is being implemented. Starting with a small project with the potential to be scaled up once influence and traction has increased, was suggested as a better way of going about practising QI.

*B.4. Training as an opportunity to promote QI*

Training was frequently discussed as a means by which to increase staff's understanding and motivation to practise QI.
